# Glycoproteome remodelling in MLL-rearranged B-cell precursor acute lymphoblastic leukemia

**DOI:** 10.1101/2021.06.25.21259296

**Authors:** Tiago Oliveira, Mingfeng Zhang, Eun Ji Joo, Hisham Abdel-Azim, Chun-Wei Chen, Lu Yang, Chih-Hsing Chou, Xi Qin, Jianjun Chen, Kathirvel Alagesan, Andreia Almeida, Francis Jacob, Nicolle H Packer, Mark von Itzstein, Nora Heisterkamp, Daniel Kolarich

## Abstract

B-cell precursor acute lymphoblastic leukemia (BCP-ALL) with mixed-lineage leukemia gene rearrangement (MLL-r) is a poor-prognosis subtype for which additional therapeutic targets are urgently needed. Currently no multi omics data set for primary MLL r patient cells exists that integrates transcriptomics, proteomics and glycomics to gain an inclusive picture of theranostic targets.

**Methods:** We have integrated transcriptomics, proteomics and glycomics to i) obtain the first inclusive picture of primary patient BCP-ALL cells and identify molecular signatures that distinguish leukemic from normal precursor B-cells and ii) better understand the benefits and limitations of the applied technologies to deliver deep molecular sequence data across major cellular biopolymers.

**Results:** MLL-r cells feature an extensive remodelling of their glycocalyx, with increased levels of Core 2-type O-glycans and complex N-glycans as well as significant changes in sialylation and fucosylation. Notably, glycosaminoglycan remodelling from chondroitin sulfate to heparan sulfate was observed. A survival screen, to determine if glycan remodelling enzymes are redundant, identified MGAT1 and NGLY1, essential components of the N-glycosylation/degradation pathway, as highly relevant within this *in vitro* screening. OGT and OGA, unique enzymes that regulate intracellular O-GlcNAcylation, were also indispensable. Transcriptomics and proteomics further identified Fes and GALNT7-mediated glycosylation as possible therapeutic targets. While there is overall good correlation between transcriptomics and proteomics data, we demonstrate that a systematic combined multi-*omics* approach delivers important diagnostic information that is missed when applying a single omics technology.

**Conclusions:** Apart from confirming well-known MLL-r BCP-ALL glycoprotein markers, our integrated multi-omics workflow discovered previously unidentified diagnostic/therapeutic protein targets.

**Graphical abstract:** 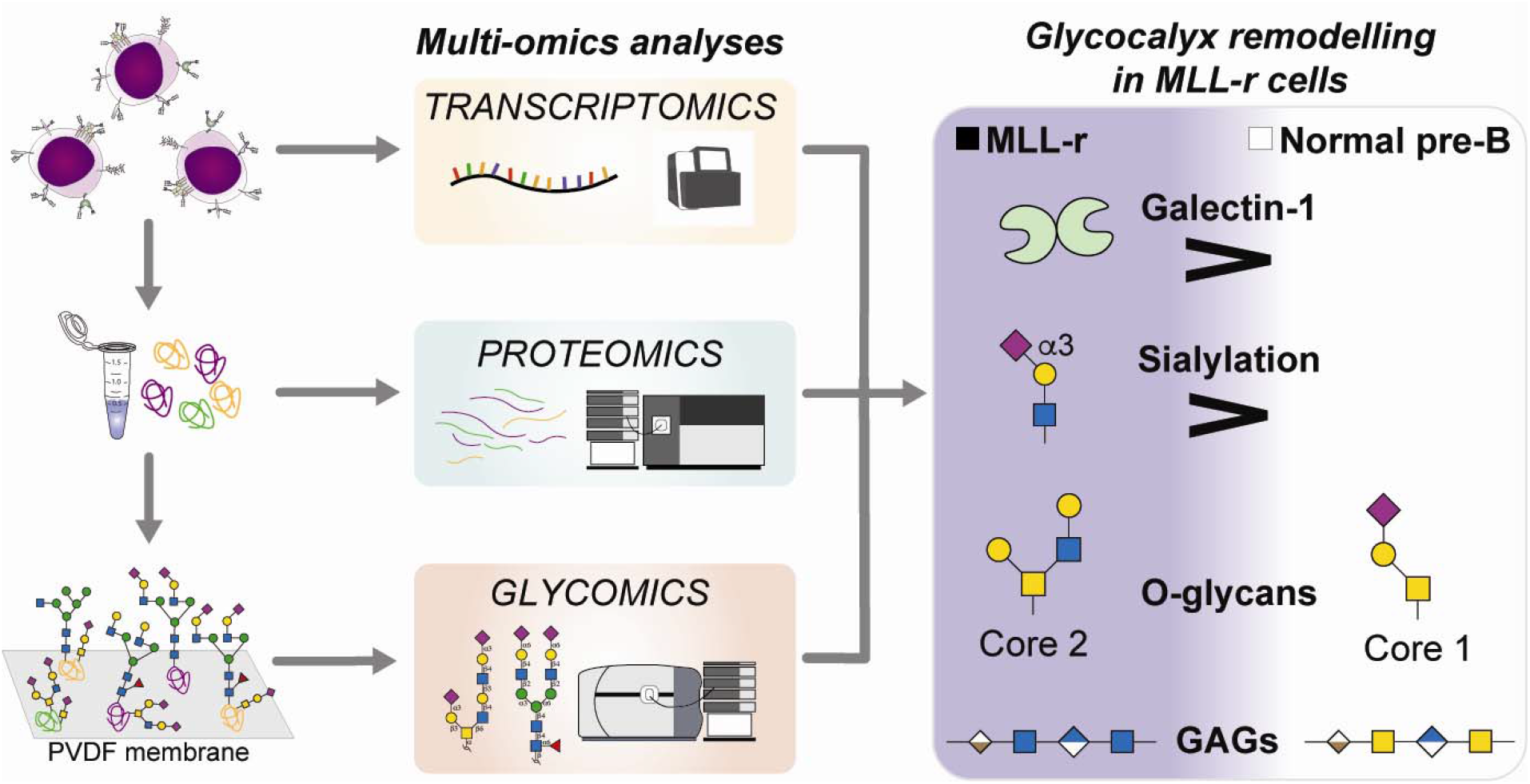

## Introduction

Each year almost half a million new patients are diagnosed with leukemia (Globocan2020) [1]. Acute leukemias including acute lymphoblastic leukemia (ALL) and acute myeloid leukemia (AML) belong to the group of more aggressive leukemias characterized by a rapid proliferation of malignant hematopoietic cells. ALL involving B-cell precursors (BCP-ALL) represents the most common type of cancer in children, and is also frequently diagnosed in adults [2]. BCP-ALL can be further subdivided into 23 categories based on molecular characteristics [3]. One subclass, MLL-r, involves rearrangements of the mixed-lineage leukemia (*MLL*) gene located on chromosome 11q23. The t(4;11)(q21;q23)/*MLL*-*AFF1*(*AF4*) is the most frequent translocation involving the *MLL* gene, but 94 other genes can also be involved [4].

More than 70 % of infants with BCP-ALL have MLL involvement. Although in general treatment options for pediatric BCP-ALL have significantly improved and overall survival rates in children exceeds 90 % [5], this specific subtype has among the lowest overall survival rates in adults as well as children [3, 6]. Furthermore, newer therapies such as the infusion of autologous CAR-T cells directed against the CD19 antigen are less effective in MLL-r B-ALL due to lineage switch [7-9]. Thus, discovering possible leukemia-specific antigens in MLL-r leukemia remains an important goal in the development of future therapeutics.

While proteins have been primarily viewed as treatment targets, it is more than likely other targets exist based on yet-to-be discovered significant cell surface differences. For example, and importantly, potential differences at the level of glycosylation as a major form of post-translational protein modification have never been explored. This is of particular significance as glycosylation affects virtually all cell surface receptors as well as the extracellular matrix, both of which are well-known to play a major role in supporting cancer cell survival [10]. Together with other glycoconjugates (proteoglycans and glycolipids), glycoproteins form the glycocalyx, a complex layer that surrounds every living cell. The glycocalyx structure is highly cell-type specific and is subject to dynamic changes, in particular as a consequence of malignant transformation [11-13]. Such cell-surface alterations are important because they impact cellular recognition processes, cell behaviour and immune responses [14-16].

However, these alterations are difficult, if not impossible, to fully determine solely using genomic approaches as they are multi-layered and are partly the outcome of genome-independent regulation. This makes a combined multi-*omics* approach the best and only option to capture a holistic and detailed picture of the glycocalyx structure. An in*-*depth multi-*omics* data set of this type, however, does not exist for primary MLL-r patient cells. In this unique study, we have undertaken such an integrated multi-*omics* investigation of primary MLL-r and control normal precursor B cells from bone-marrow. As outlined in the workflow shown in *Suppl. Fig. 1*, we have undertaken the first combined transcriptomics study that incorporates both glycomics and proteomics analyses for MML-r primary patient cells. Our results provide the first reported evidence that MLL-r cells have undergone a radical transformation of the protein glycocalyx when compared with healthy precursor B-cells. These data provide an exciting advance towards the development of novel therapies targeting this low-survival leukemia subtype.

## Methods

### Reagents

Water was purified using a Milli Q-8 system (Merck KGaA, Darmstadt, Germany). High quality-grade reagents were purchased from Sigma-Aldrich (St. Louis, MO, USA), unless otherwise mentioned. PNGase F (Cat#P0704) was from New England Biolabs, 500,000 U/mL. Sequencing-grade Trypsin was from Roche (Cat#11047841001). High-grade chloroform and methanol used to perform protein precipitation were from Merck (Cat#1024442500 and 1060072500, respectively).

### Ethics statement

All human specimen collection protocols were reviewed and approved Institution Review Boards. All methods were performed in accordance with the relevant guidelines and regulations. Collections were in compliance with ethical practices and Institution Review Boards approvals.

### Pilot glycan isolation study

Primary leukemia samples typically undergo some processing. To determine if such procedures would significantly affect glycan recovery, we tested this on biological duplicate samples from the MLL-r B-cell precursor ALL cell line RS4;11 (ATCC #CRL-1873; established from patient Bone Marrow (BM) leukemia cells with an MLL-AF4 fusion protein). We compared RBC lysis, Ficoll density gradient /mononuclear cells isolation, flow sorting, and 10 % DMSO freezing medium/freeze/thaw and samples receiving no further treatment (*Suppl. Fig. 2A*). In comparison, we also included samples grown on OP9 stromal cells. *Suppl. Fig. 2B* shows that relative intensities of recovered glycans were comparable between different procedures. Our analytical approach also allows discrimination between closely related sialylated glycan structures (*Suppl. Fig. 2C*).

### MLL-r and normal control precursor B cell isolation

Starting materials for isolation of control healthy precursor B cells were four BM samples (R7B1, R7B3, R7B6 and R7B11) from different donors depleted of CD34+ stem cells. R7B11 was not further processed, whereas R7B6 was enriched in CD19+ B cells, and R7B1 and R7B3 were enriched for both B- and T-cells [CD19 and TCRα/β]. To isolate normal CD19+CD10+ precursor B-cells, around 2×10^9^ of such viably frozen normal bone marrow cells from each sample were applied to EasySep™ Release Human CD19 Positive Selection Kit (STEMCELL Technologies, Vancouver, BC, Canada, Cat#17754) and EasySep™ Human CD10 Positive Selection Kit (STEMCELL Technologies, Cat#18358) columns.

MLL samples in this study were BM0 [(11q23) relapse sample, 98 % blasts], BM37 [(46XY, t(9;11) diagnosis, 98 % blasts], and BM41 (60 % blasts, MLL rearranged (11q23; t9;11) 46XY]. Ficoll-Plaque Plus (GE Healthcare, Cat#17-1440-02) centrifugation was used to remove red blood cells, according to manufacturer’s instructions and after washing viably frozen with DMSO. Viably frozen cells were thawed and washed twice with ice-cold DPBS (500*xg*, 4 min, 4 °C). Each sample, consisting of 4-6×10^6^ total cells, was divided into two fractions, one for proteomic/glycomic analyses and the other for RNA sequencing.

### Sample preparation for (glyco)protein extraction

Frozen cell pellets of normal bone marrow (n=4) and MLL-r (n=3) cells were processed according to the same protocol: 2-3 million cells per sample were lysed in cell lysis buffer composed of Dulbecco’s PBS (Sigma, Cat#D8537) containing 1 % Triton X-100 (Sigma, Cat#T8787) and Protease Inhibitors (Sigma, Cat#P8340). Briefly, 400 µL of the cell lysis buffer were added to each sample, and tubes were kept on ice for 20 min. Samples were then homogenised using a T-10 ULTRA-TURRAX® (IKA, Cat#0003737000) in 3 × 3 sec cycles, followed by 30 sec on ice between cycles. Samples were kept on ice for 10 more minutes, after which sonication was performed for 30 sec using a water-bath sonicator to shred DNA. Samples were centrifuged for 15 min at 15,000*xg*, 4 °C. Supernatants were collected to new tubes and all resulting aliquots were stored at -80 °C prior use. After lysis, all samples were quantified in triplicates using a Pierce™ BCA Protein Assay Kit (Thermo Scientific, Cat#23225).

Protein content extracted from similar number of cells from samples R7B11 and BM41 was on average approximately 25 µg compared to 115 µg or more for the other 5 samples (*Suppl. Table 1*). To ensure high-quality proteomics and glycomics preparations, these samples were excluded from the final preparations. The protein lysates of the other five samples were used as described in each individual method and as presented in *Suppl. Table 1*.

### RNA seq and RNA expression analysis

RNA was extracted from frozen cell pellets using Trizol (Therm*O-*Fisher 315596026) and further purified using a RNeasy Mini kit (Qiagen #74104). Quality control analysis was done using Bioanalyzer RNA Nano and D1000Chips. Nucleic acid concentrations were determined by Qubit. The mRNA library was prepared using an Illumina Truseq Stranded mRNA High Throughput Prep kit and samples were sequenced using an Illumina NextSeq 500 Mid-Output Sequencing Reagent kit (v2, 150 cycles), 132 M reads on an Illumina NextSeq 500 instrument. RNA-seq results were aligned twice to the human genome. One analysis used the GRCh37 annotation file (including around 50,000 genes including pseudogenes and non*-*coding RNA) whereas the second alignment, and of which results were used for proteomic comparisons presented here, made use of the genome assembly GRCh38.p13 (GCA_000001405.28) genome annotation files (around 19,862 mainly protein*-*encoding genes but not including IGHM). For the latter analysis, reads were quality-checked and processed using the nf-core/rnaseq analysis workflow v1.3 pipeline consisting of Nextflow v19.04.1, FastQC v0.11.8 Cutadapt v2.4, Trim Galore! v0.5.0, STAR vSTAR_2.6.1d, HISAT2 v2.1.0, Picard MarkDuplicates v2.18.27, Samtools v1.9, featureCounts v1.6.4, StringTie v1.3.5, Preseq v2.0.3, deepTools, v3.2.0, RSeQC v3.0.0, MultiQCv1.7 software. Raw count results were analysed using edgeR to determine significantly regulated genes (criteria: fold change ≥2; p<0.05; low expression filter rpkm <1.0). There were 3936 genes meeting the default criteria which were differentially expressed in this comparison, with 1883 genes up-regulated and 2053 genes down*-*regulated. The normalized RNA counts were plotted using GraphPad Prism (v8.4.3). Diagnosis leukemia samples including 70 MLL-r cases and 74 normal bone marrow controls [17] from the MiLE study (GSE13159) were compared for expression of GAG synthesis enzymes using online tools [18].

### N*-* and O-glycan release for glycomics analyses

50 µg of (glyco)proteins were reduced by adding a volume of 500 mM of Dithiothreitol to each sample, to a final concentration of 20 mM, at 50 °C for 1 hour. After cooling, samples were then subjected to alkylation using 40 mM Iodoacetamide in the dark at room temperature. The reaction was quenched by adding another 20 mM of Dithiothreitol and incubating for 5 minutes.

Proteins were precipitated using the Chloroform:Methanol:Water separation as described previously [19]. The resulting protein pellet was left to air dry for 10 minutes. 5 µL of 8 M urea were added to each sample to resuspend the protein pellet using intensive vortexing. The final concentration of urea was adjusted to 4 M by adding 5 µL of pure water (MQ-H_2_O).

The urea dissolved proteins were dot blotted onto a PVDF membrane (Immobilon*-*P, 0.44 µm pore, Merck Millipore), and N*-* and O-glycans were released as described previously [20] (see also *Supplementary Methods* for details). Before mass spectrometry analyses, N*-* and O-glycans were carbon cleaned off-line using porous-graphitised carbon (PGC) material packed on top of C18 ZipTips to avoid any potential contaminants and then stored at -20 °C until their PGC-LC-ESI-MS/MS analyses.

### PGC-nanoLC-ESI-MS/MS glycomics

The N*-* and O-glycome was determined using the PGC-nanoLC-ESI-MS/MS glycomics technology as described previously [20-22] (see also *Supplementary Methods* for details). Previous studies relating to glycosylation and BCP-ALL focused on the identification of particular glycoconjugates or glycan traits, such as the acetylation of sialic acid, 9-*O-*acetyl-Neu5Ac [23, 24]. We note that the technologies used here do not allow to routinely evaluate the level of *O-*acetylation of sialic acids, as these labile modifications are lost due to the buffers used during the PGC-LC-ESI-MS/MS analyses. In addition, because of limited cell numbers, this study did not examine the samples for GAGs. All details are described following the respective MIRAGE (Minimum Information Required for A Glycomics Experiment) guidelines in the supplementary Material [25-28].

### Glycan structure determination and relative quantitation

Glycan structures were determined as previously described [21, 22] (see also *Supplementary Methods* for details). Unsupervised clustering analysis of the relative glycan abundances, and the respective heat map representation, were performed using the package *pheatmap* (v1.0.12) available in R studio (v1.3.1073). The relative intensities were also plotted using GraphPad Prism (v8.4.3), and *p-*values were calculated by performing an unpaired *t*-test. Symbols of calculated significance (*p*<0.01, **\***) are represented when groups are significantly different. All represented N- and O-glycan structures and monosaccharides are depicted following the rules of the Symbol Nomenclature for Glycans (SNFG) [29, 30].

### Cas9-CRISPR screen of glyco*-*enzymes

KOPN8 (https://web.expasy.org/cellosaurus/CVCL_1866), an MLL-r B-cell precursor ALL cell line, was genotype-verified using STR genotyping. Cells were stably transduced with a lentiviral vector containing a blasticidin-selection marker and expressing the Cas9 protein [AddGene #52962 plasmid [31]]. The sgRNA LV library was constructed in the pU6-sgRNA-EF1Alpha-PURO-T2A-RFP (ipUSEPR) vector [32]. Each neutral selection gene control [neg, LacZ, Luc and Ren] was covered by ten sgRNAs each and 2 sgRNAs each were directed against ten essential gene controls [PCNA, POLR2D, POLR2A, PRL9, RPL2, CDK9 RPA3, RPS20, MYC, BRD4]. Cells transduced with LV expressing the latter sgRNAs would be expected to be depleted from the cell population. Target glycogenes were covered by 10 sgRNAs each. The entire screen included 1082 sgRNAs with 102 genes encoding glycan-remodelling enzymes, 4 neutral genes and 11 genes of which ablation is expected to reduce cell growth and viability. On d0 biological duplicates of around 20×10^6^ KOPN8/Cas9 cells in 1640 medium were transduced using 10 µg/mL polybrene at a low MOI to ensure that most cells would be transduced with one or no sgRNAs. After 24 hrs, puromycin selection at 4 µg/mL was applied for 9 days, and 3 µg/mL puromycin was used from d10-d32. On d20 of selection almost all cells contained an ipUSEPR construct based on FACS for the RFP marker. Cells were harvested for DNA isolation on d28. After d32 cells were also plated on an OP9 stromal feeder layer and grown for an additional period under puromycin selection. Cells in the culture supernatant were harvested on d48 and also used for DNA isolation to obtain two biological replicates. Each isolate from 1-5×10^6^ cells contained sufficient DNA to yield an approximately 1000x coverage. DNAs were sequenced on an Illumina NextSeq 550. Results were analyzed using MAGeCK [Model-based Analysis of Genome-wide CRISPR-Cas9 Knockout [33] https://hpc.nih.gov/apps/MAGeCK.html. GiniIndex values for numbers of sgRNAs with 0 read counts varied between 0.09 and 0.36. The median-normalized read counts and the distribution of read counts were comparable across samples.

### High-pH fractionation and C18-nanoLC-ESI-MS/MS analyses of peptides

50 µg of protein (samples R7B1, R7B3, R7B6, BM0 and BM37) and 10 µg (samples R7B11 and BM41) were reduced, alkylated and precipitated with Chloroform-Methanol as described above. The protein pellet was air-dried for 10 minutes before 100 µL of 25 mM of ammonium bicarbonate (Sigma, Cat#09830) were added to the pellets. Trypsin was added at a ratio 1:25 (enzyme:protein ratio) and samples were incubated for 18 hours at 37 °C. After digestion, trypsin was heat-inactivated at 95 °C for 10 minutes, and samples were dried under vacuum. 1000 U of PNGase F (2 µL) prepared in 50 µL of H_2_^18^O (Sigma, Cat#329878) were added, and samples were incubated at 37 °C for 3 hours to deglycosylate N-linked glycopeptides before drying under vacuum.

Peptides were resuspended in 300 µL of 0.1 % trifluoroacetic acid (TFA) and fractionated using a Pierce™ High pH Reversed-Phase Peptide Fractionation Kit (Sigma, Cat#84868) following the manufacturer’s instructions. Briefly, the resuspended peptides were loaded to the pre-conditioned supplied spin columns, and washed (3,000*xg*, 2 min) once using water. Increasing concentrations of acetonitrile (ACN) (5 %, 7.5 %, 10 %, 12.5 %, 15 %, 17.5 %, 20 %, and 50 %) in 0.1 % triethylamine (TEA) buffer were used to elute (3,000*xg*, 2 min) the bound peptides into eight distinct fractions. All the resulting fractions were dried under vacuum and kept at -20 °C until analysis. Samples were resuspended in 0.1 % TFA and peptide amounts were quantified using a Thermo Scientific™ NanoDrop™ One/OneC Microvolume UV-Vis Spectrophotometer.

The off-line fractionated peptides were identified using an Orbitrap Fusion™ Tribrid™ Mass Spectrometer coupled to an UltiMate™ 3000 UHPLC nanoLC (both Thermo Scientific™) (*Supplementary Methods* for details on the columns and methods used). Based on the NanoDrop quantitation, a volume corresponding to 600 ng of peptides were injected of each fraction.

### Proteomics data analyses

All files were analysed using the Andromeda search engine integrated into the MaxQuant suit (v6.10.43) [34]. For high pH fractionated sample analyses using MaxQuant, the 8 fractions were combined according to their respective sample. As two injections were made for each fraction, this resulted in two combinations of 8 fractions, namely injection 1 and injection 2 for each sample. The HCD-MS/MS spectra were searched against *in silico* tryptic digest of *Homo sapiens* proteins from the UniProt sequence database (v10; May 2020) containing 20,359 protein sequences (Swiss-Prot IDs). All MS/MS spectra were searched with the pre-set MaxQuant parameters, and the following modifications were used: cysteine carbamidomethylation was set as a fixed modification; methionine oxidation, acetylation of protein *N-*terminus, and asparagine deamination and ^18^O deamination were allowed as variable modifications. False discovery rate (FDR) of the peptide spectral matches (PSMs), protein, and site were set to 1 % based on Andromeda score. Match between runs (MBR) algorithm was activated to allow matching MS features between the different sample fractions and improve quantification [34].

LFQ-Analyst was used for the label-free quantitation (LFQ) of the MaxQuant pre-processed proteomic datasets [35]. Two main “Conditions” were defined as “MLL-r” and “Normal BM”, and each injection was used as an independent replicate. In the Advanced Options setting, the “Adjusted *p*-value cut-off” was defined to 0.01 (q-value, FDR<1 %), whilst the “Log_2_ fold change cut-off” (log_2_FC) was defined to 2.

The log_2_ and *p*-values calculated by LFQ-Analyst were used to generate a volcano plot representation using the package *EnhancedVolcano* (v1.7.16) in R studio (v1.3.1073). GraphPad Prism (v8.4.3) was used for the RNA-protein integrative analyses, by plotting the calculated magnitude (log_2_) differences derived from our proteomics and RNA-seq analyses, targeting solely the differentially expressed proteins.

## Results

### MLL-r patient cells undergo a distinct protein O*-*glycome transformation

The initiating step of protein O*-*glycosylation is tightly controlled by 20 distinct GALNTs enzymes that post-translationally transfer a *N*-acetylgalactosamine (GalNAc) on folded glycoproteins [36]. Of these, GALNT1, 2 and 3 are considered to be the most widely expressed and responsible for glycosylating the bulk of glycoprotein acceptor substrates [37]. Each GALNT has specific preferences for the protein sequence/structure motifs that it can glycosylate, and the activity of some GALNTs can also depend on the previous action of other GALNTs. Because these enzymes are highly regulated in a cell-, tissue- and protein*-*specific manner [37, 38], O*-*glycosylation is a major regulator of cell function [16]. RNA-seq analyses identified ten *GALNT* gene transcripts of which six showed expression level differences between MLL-r and control cells (*Fig. 1A, Suppl. Table 2*). On a protein level, expression of GALNT1, GALNT2 and GALNT7 was confirmed, which also revealed significantly increased GALNT7 in MLL-r cells (*Fig. 1B* and *Suppl. Table 3*).

**Figure 1.**
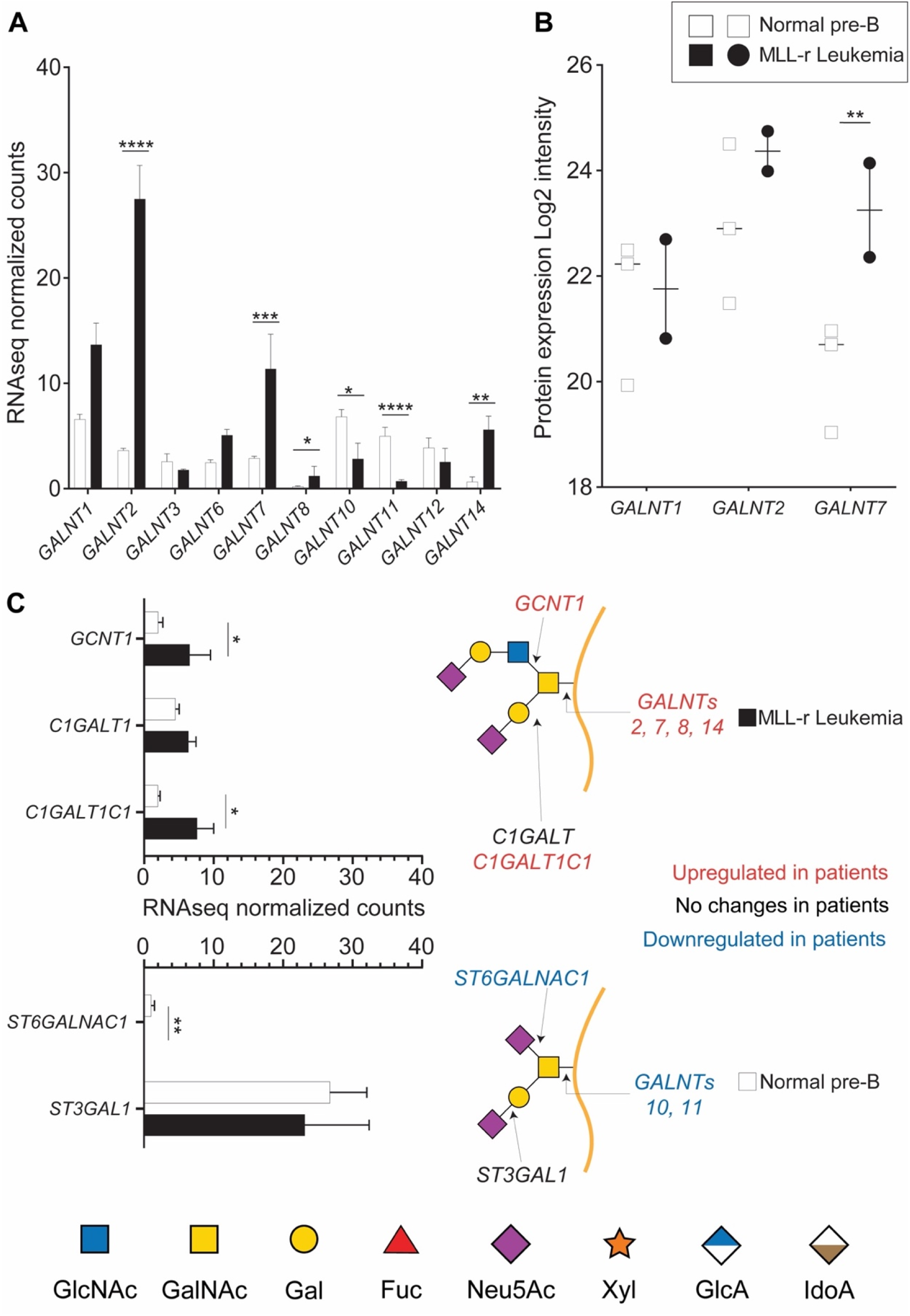
MLL-r cells have altered expression of key enzymes involved in O*-*glycan synthesis. RNA-seq (A, C) or proteomic (B) results for the indicated enzymes. Symbols in panel (B) each represent one sample. Panel (C) also shows a schematic structure of two glycans and the enzymes responsible for their synthesis. *p<0.05; **p<0.01; ***p,0.001; ****p<0.0001

The activity of GALNTs is the rate-limiting first step in O*-*glycan biosynthesis, but further O*-*glycan extension is regulated by the concerted and competitive action of various glycosyltransferases (GTs). The mRNA expression levels of GTs responsible for extending the 3-position of the initial GalNAc residue, such as C1GALT1 (encoded by *C1GALT1*), were unchanged. In contrast, expression levels of its essential molecular chaperone Cosmc (*C1GALT1C1*) [39, 40] doubled in MLL-r cells (*Fig. 1C*). mRNA expression was three-fold increased for *GCNT1*, the transferase responsible for initiating Core 2 type O*-*glycan synthesis (*Fig. 1C*), whereas expression of *ST3GAL1*, the sialyltransferase known to add a sialic acid on the core 1 galactose, remained unchanged (*Fig. 1C*). In addition, expression of *ST6GALNAC1*, the sialyltransferase competing with GCNT1 and thus preventing Core 2 type O*-*glycosylation, was very low in MLL-r cells (*Fig. 1C*). These transcriptomics data suggest a major remodelling of the MLL-r O*-*glycome.

O*-*glycomics by LC MS/MS confirmed that MLL-r BCP-ALL cells exhibited a significantly altered O*-*glycome, shifting towards Core 2-type O*-*glycans, while Core 1-types were the major forms in normal control BCP cells. Overall, we identified 21 distinct O*-*glycans, including five Core 1, thirteen Core 2 and two O*-*fucose type glycans next to a sialylated hexose disaccharide (*Fig. 2A* and *2B*; *Suppl. Table 4*). In MLL-r cells, 51 % of all glycans were Core 2 O*-*glycans compared to 26% in normal BCP cells. The level of Core 1 type O*-*glycans was almost halved in MLL-r (37 % *versus* 64 %, *Fig. 2B* and *C*).

**Figure 2.**
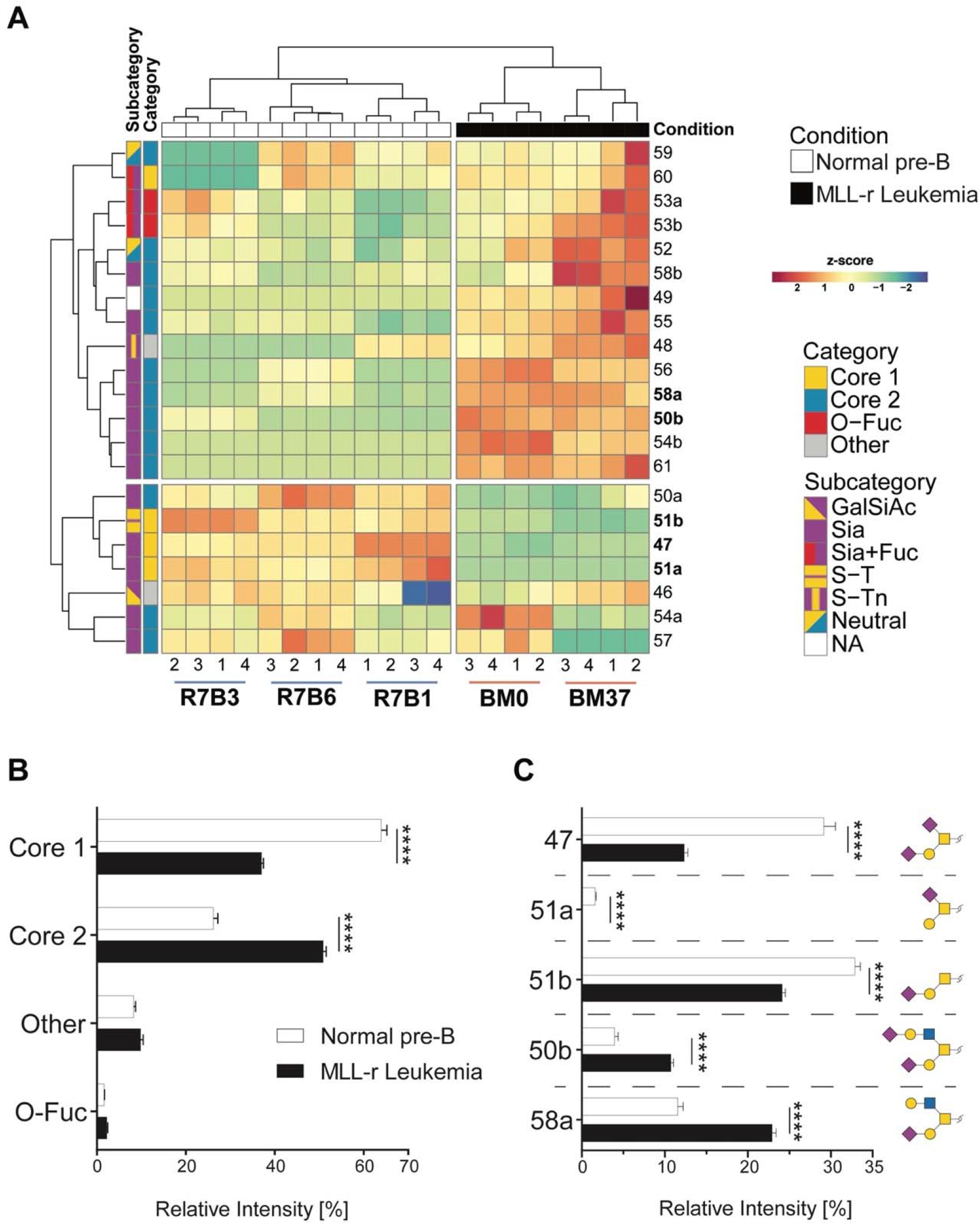
The O-glycome differs between MLL-r and normal BCP cells. (A) Heat map showing the z-score of the detected relative abundance of the O-glycan structures indicated with a number to the right (see *Suppl. Table 4* for corresponding structures). Sample identifiers are indicated below the image. Numbers refer to technical replicates. Glycans are categorized as indicated in the key to the right. (B) Relative intensity (as percentage of the total) of the major categories of O-glycans found in MLL-r and normal BCP cells. (C) Representation of the five most abundant and important sialylated O-glycan structures. Monosaccharide symbols are represented according to SNFG guidelines [29, 30]. ****p<0.0001

More than 1300 human proteins are reported to be O*-*glycosylated [41-43]. We found that mRNAs for around 70 % of these were expressed in MLL-r/control cells, with 40 % exhibiting significantly different expression in MLL-r samples (159 higher, 197 lower, *Suppl. Table 2*) compared to normal precursor B cell controls. Proteomics confirmed the presence of 241 previously reported O*-*glycoproteins, of which 33 were differentially expressed (with 9 upregulated in MLL-r cells; *Suppl. Table 3*). Together, these glycomic, transcriptomic and proteomic data provide clear evidence for the extensive remodelling in the O*-*glycoprotein and O*-*glycan components of the MLL-r cell glycocalyx.

### Sialylation and Lewis X fucosylation are increased on N*-*glycans of MLL-r cells

Protein N*-*glycosylation is critical for correct protein folding, cell-cell recognition and cellular interactions [44]. In both MLL-r and control cells, oligomannose and complex type N*-*glycans were the main forms of N*-*glycosylation (*Fig. 3A*). MLL-r cells showed increased complex type N*-*glycan levels compared to controls (52.4 % *versus* 42.6 %), while the levels of paucimannose (10.9 % and 11.9 %) and hybrid type (≈5 %) N-glycans were similar. The complex type N*-*glycans were mainly biantennary. Only 5 % of all N-glycans showed features consistent with tri- or tetra-antennary structures, but these could not be further characterized as a consequence of their low abundance levels.

**Figure 3.**
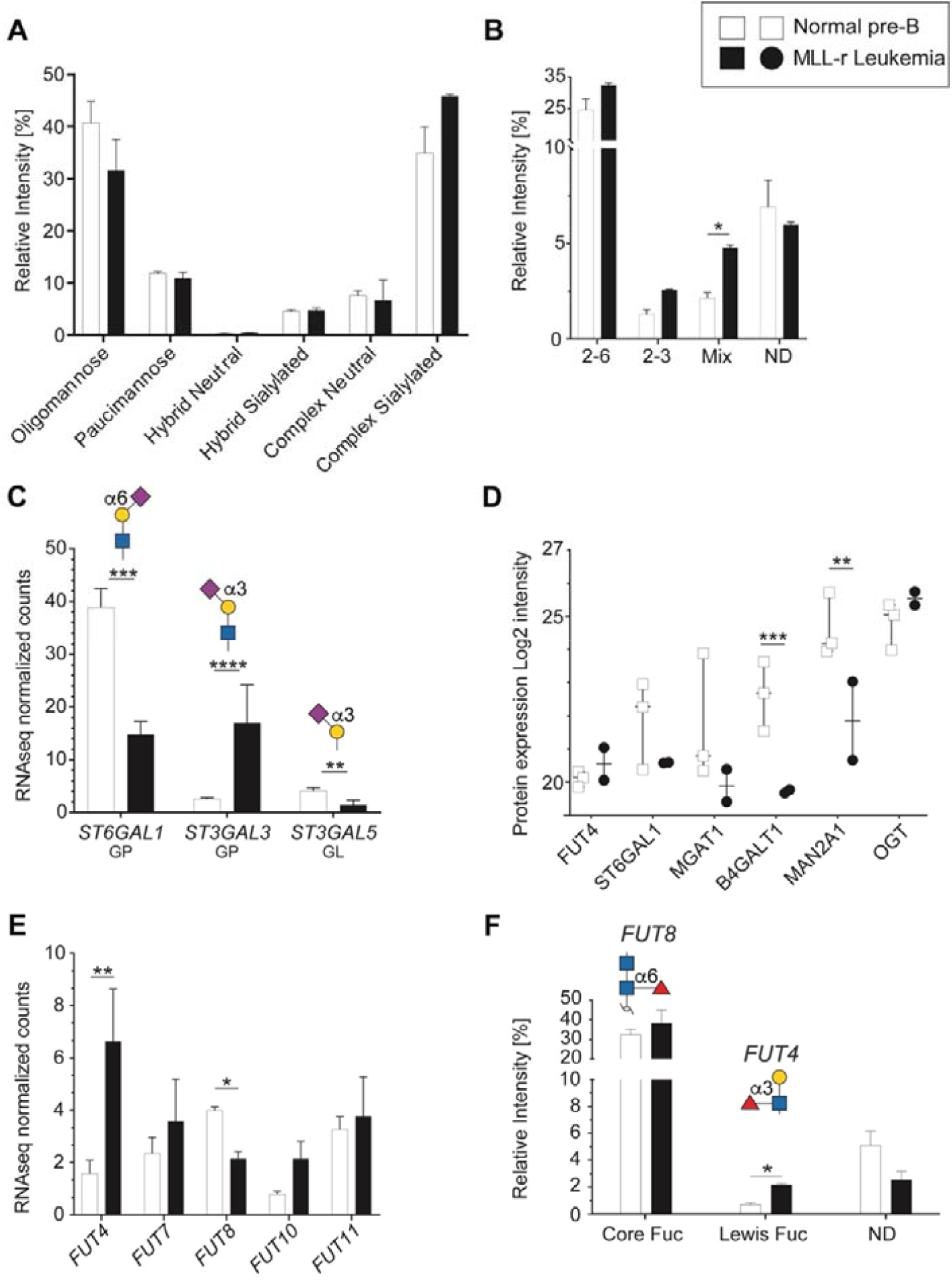
Differential sialylation and fucosylation of N-glycans in MLL-r B-ALL samples. (A) Relative intensities of total N-glycans indicated in the graph, as detailed in *Suppl. Table 5*. (B) Relative intensity of different sialic acid linkages on N-glycans. MLL-r cells contained higher levels of tri-antennary, fully sialylated N*-*glycans (*Suppl. Table 5*, structures 43a and b) while incompletely sialylated, tri-antennary and also tetra-antennary N*-*glycans (41b and 42a, respectively) were the most abundant forms of these branched N*-*glycans found on the controls. (C) Normalized mRNA expression of the indicated sialyltransferases, and their respective resulting glycan epitope products as shown in the graph. (D) Protein expression levels as log2 intensity of the indicated glycosyltransferases. (E) Fucosylated glycans present in MLL-r and control samples. The structures and the fucosyltransferases responsible for their synthesis are shown above the graph. (F) Fucosyltransferase RNA expression. *p<0.05; **p<0.01; ***p,0.001; ****p<0.0001

The overall N*-*glycan sialylation levels were increased in MLL-r cells, with N*-*glycan structures carrying α2-6, both α2-3 and α2-6, or exclusively α2-3 linked sialic acids (*Fig. 3B*). These glycomic findings correlated well with the increased *ST3GAL3* mRNA levels (*Fig. 3C*), which is likely to be the cause of the observed increased sialylation of tri- and tetra-antennary N*-*glycans in MLL-r cells. For ST6GAL1, however, a correlation between the decreased mRNA and protein levels in MLL-r cells and a decrease in α2-6 sialylation could not be found, as the overall α2-6 sialylation levels remained unchanged (*Fig. 3B-D*).

The functional behaviour of glycoproteins and cells is also well-known to be influenced by fucosylation [45, 46]. The human genome contains 13 functionally-distinct fucosyltransferase genes (*FUTs*), of which 5 were detected at the transcript level (*Fig. 3E*). Our glycomics analyses showed that core fucosylation, a product of FUT8 activity, was the major fucose modification present on about one-third of all N-glycans. While *FUT8* transcript levels were lower in MLL-r, no difference in core-fucosylation levels on the glycans was observed between MLL-r and control cells (*Fig. 3E* and *F, Suppl. Table 5*). In contrast, Lewis X type fucose was present on less than 1 % of N*-*glycans in controls, but was almost triple the level in the MLL-r cells (*Fig. 3F*). Consistent with the glycomics data, transcripts for *FUT4*, one enzyme responsible for Lewis X synthesis [47, 48], were increased more than threefold (*Fig. 3E*). Overall, these data indicate a slight shift in the MLL-r N*-* glycome towards more complex type and Lewis X fucosylated N*-*glycans. Importantly, our data also indicate that a change in glycosyltransferase transcript levels is not automatically reflected in a change in the actual protein glycosylation.

### Terminal lactosamine levels are increased in MLL-r

The protein levels of B4GALT1, the enzyme responsible for transferring galactose to generate the LacNAc epitope (Galβ1-4GlcNAc-) [47], were reduced 2.9-fold in MLL-r cells (*Fig. 3D*; *Suppl. Table 5*), without observing corresponding changes in *B4GALT1* mRNA levels (*Suppl. Table 2)*. Interestingly, terminal (non*-*reducing end) LacNAc epitope levels on both N*-* and O-glycans were significantly increased in MLL-r cells. This included LacNAc glycoepitopes capped with α2-3 NeuAc residues (*Fig. 4A* and *4B*) that have recently been confirmed to be recognised by Galectin*-*1 and -3, while α2-6 sialylation of their counterparts prevents Galectin binding [49]. Galectins are glycan*-*binding proteins involved in key physiological processes such as inflammation and signalling [50-52]. They are also critical components of the tumour microenvironment, in particular in the bone marrow and in the context of various forms of leukemia [53]. An increase of Galectin*-*1 has previously been reported in BCP-ALL, and specifically in the MLL-r subtype of ALL [54, 55]. This is also supported by our data that showed an increase in Galectin*-*1 (*LGALS1*) mRNA and protein levels in MLL-r cells (*Fig. 4C* and *4D*, respectively). The transcript levels of Galectin*-*3 (*LGALS3*) and Galectin*-*3 binding protein (*LGALS3BP*), important regulators of innate immune responses often found upregulated in various cancer types [56], were significantly decreased in MLL-r cells. Reduced LGALS3BP was also confirmed at the protein level (*Fig. 4C* and *D*). Thus, the observed glycocalyx differences between MLL-r and normal bone marrow (NBM) control cells are expected to significantly impact their ability to interact with and be recognised by glycan*-*binding proteins.

**Figure 4.**
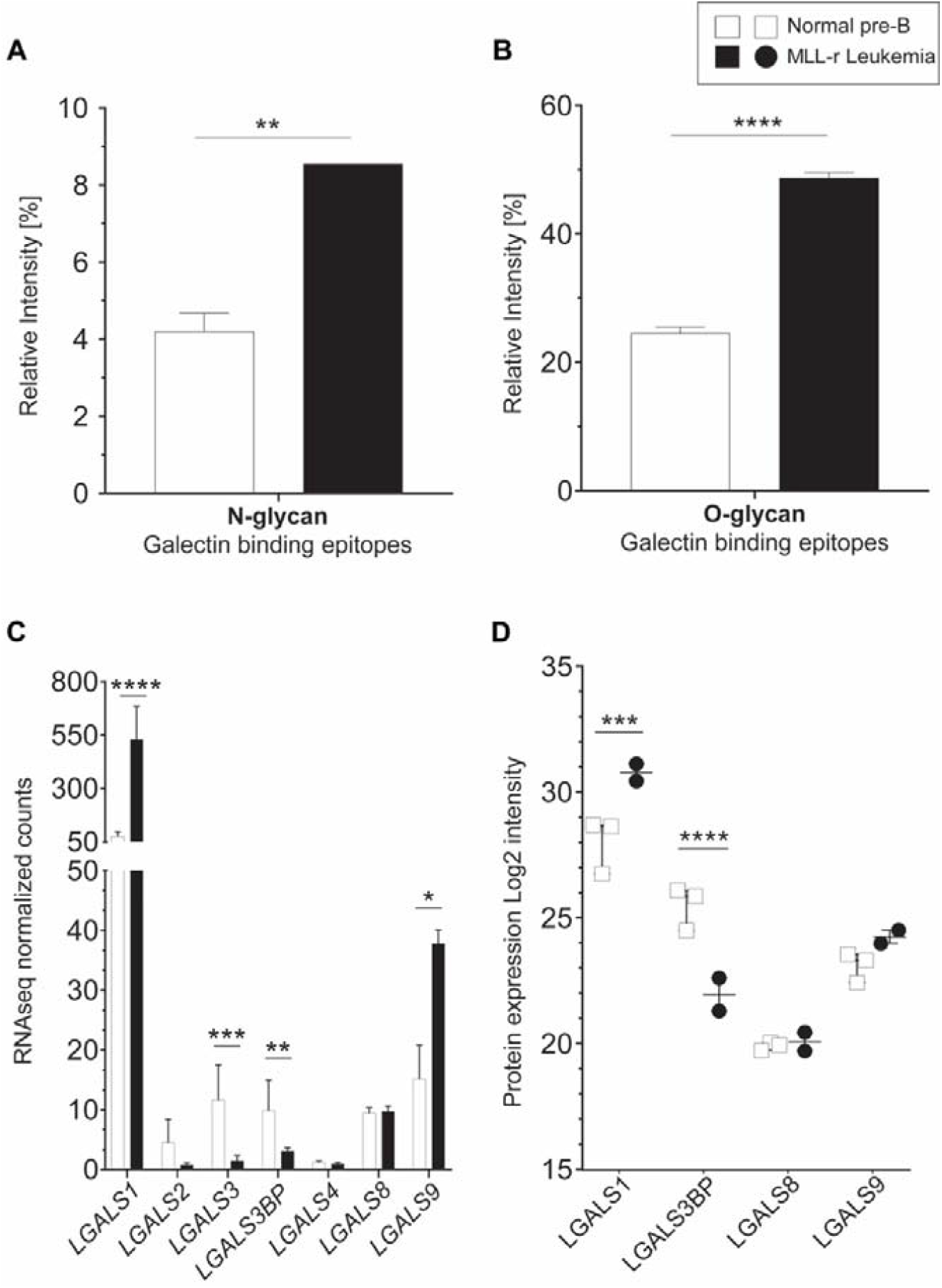
Increased terminal lactosamine in MLL-r samples as ligands for Galectin*-*1. (A, B) Relative abundance of N-glycans (A) and O-glycans (B) containing terminal lactosamine. Binding epitopes are defined as LacNAc or α2-3NeuAc-LacNAc structures. (C, D) Galectin mRNA and protein levels. *LGALS1* (Galectin*-*1) mRNA (C) and protein (D) are overexpressed in MLL-r samples compared to normal controls. *p<0.05; **p<0.01; ***p,0.001; ****p<0.0001

### Combined proteomics and transcriptomics suggest remodelling of the MLL-r proteoglycome

We have undertaken a quantitative proteomic and RNA-seq analyses of GAG-associated transferases as proxies for potential glycocalyx changes affecting proteoglycans, due to the limited availability of patient-derived primary cell material. As shown in *Fig. 5A (right panel, GAG linker)*, heparan sulfate (HS) and chondroitin sulfate (CS) share a common tetrasaccharide linker (-GlcAβ3Galβ3Galβ4Xylβ-Ser/Thr) that is attached via a xylose residue to a Ser or Thr residue [57]. In MLL-r cells, two enzymes involved in the GAG linker biosynthesis, *B3GALT6* and *XYLT1*, showed increased mRNA expression *(Fig. 5A right panel, GAG linker*, and *Suppl. Fig. 3*).

**Figure 5.**
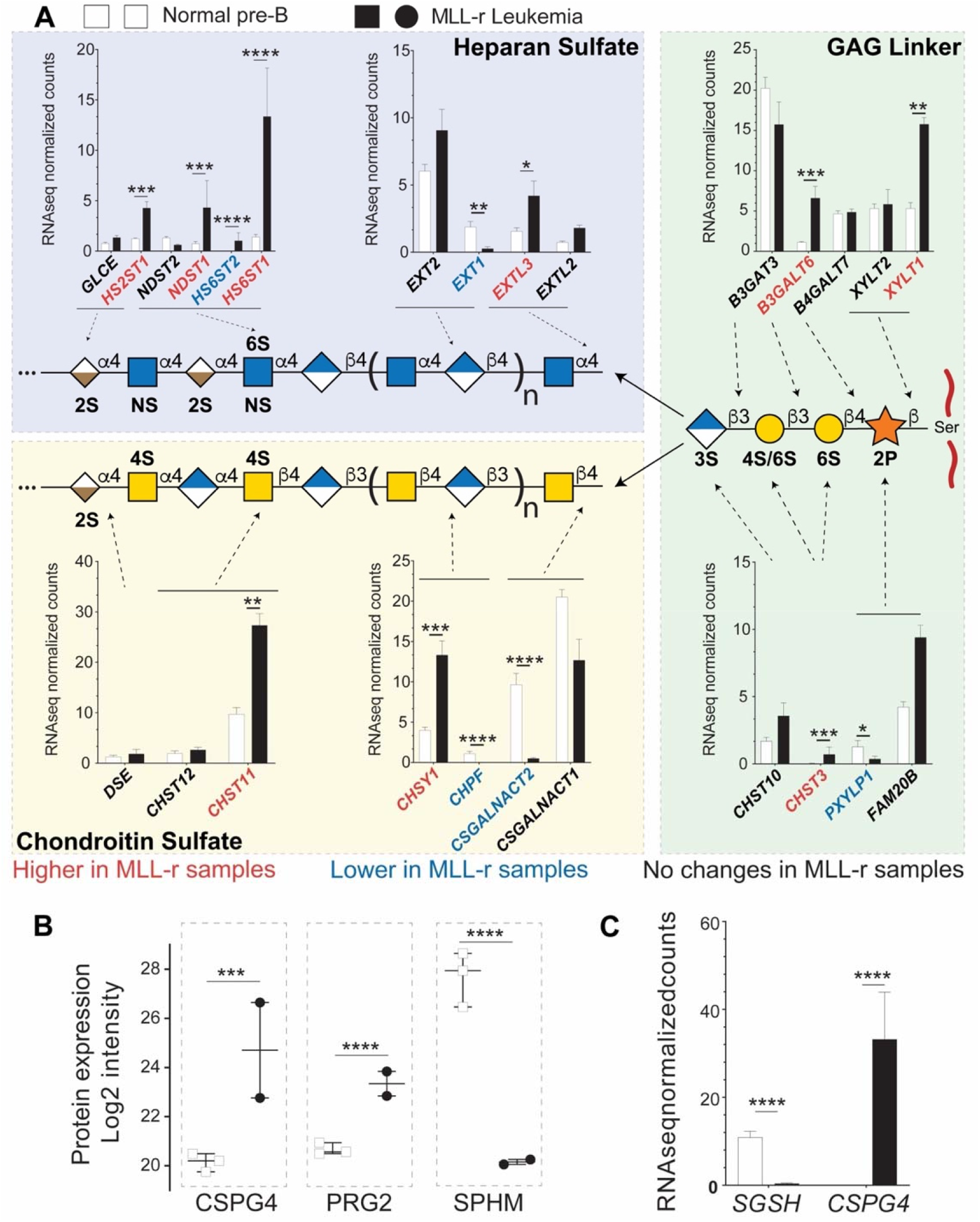
Combined proteomic/transcriptomic analysis of MLL-r samples indicates a shift toward increased HS proteoglycan glycosylation. (A) Relative expression levels of mRNAs for the enzymes involved in the synthesis of the GAG linker are shown, with red/blue depicting higher and lower levels in the MLL-r compared to control samples. The mRNA expression of enzymes involved in GAG chain elongation including mutually exclusive heparan sulfate or chondroitin sulfate glycosylation is represented. Five different transferases are involved in synthesising this tetrasaccharide, before it is elongated either as an HS or a CS chain. (B, C) Protein and mRNAs for proteoglycans differentially expressed in MLL-r samples. 11 peptide spectral matches from 2 unique peptides (in MLL-r only) were identified for PRG2. *p<0.05; **p<0.01; ***p,0.001; ****p<0.0001

This linker tetrasaccharide is the starting point to synthesise either CS or HS-chains. In MLL-r cells, *CSGALNACT1* and *CSGALNACT2* transcripts were reduced while *EXTL2* and *EXTL3* transcripts showed an increase (*Fig. 5A, left panels)*. These data suggest that the GAG glycocalyx of MLL-r cells undergoes a major remodelling compared to control cells with a shift from CS towards HS (see also *Suppl. Fig. 3*). This shift is further supported by the fact that mRNA and protein expression levels of *N-*sulfoglucosamine sulfohydrolase (*SGSH*/SPHM, *Fig. 5B, Suppl. Fig. B*), a hydrolase involved in HS degradation, are significantly decreased in MLL-r cells. On the other hand, significantly increased levels of bone marrow proteoglycan (PRG2), a potent inhibitor of heparanase (HPSE) [58] and also a proteinase inhibitor [59], were unambiguously identified at the protein level (*Fig. 5B*) even though transcript levels were below the cut-off value of <1 rpkm. HPSE itself was not detected at the protein level, and no significant trends were observed in terms of the expression of the *HPSE* transcript (*Suppl. Table 2*). These data clearly suggest (1) the presence of HS and (2) the active protection of HS in MLL-r cells, and suggest the proteoglycome as novel potential targets for MLL-r, and support the notion that the proteoglycome presents novel potential therapeutic targets for MLL-r.

### CRISPR screen supports redundancy in the critical role of glycan*-*remodelling enzymes on cell survival

Taken together, a dramatic remodelling of the glycome of MLL-r leukemia cells compared to normal BCP cells is apparent. We next selected 102 genes encoding glyco-enzymes expressed in MLL-r cells to determine if any are uniquely critical to cell survival or if that functional redundancy is likely to exist. We performed a CRISPR library dropout screen in which each gene was represented by 10 different sgRNAs (*Suppl. Table 6)*. In this type of assay, sgRNAs are introduced into cells via lentiviral transduction at a very low multiplicity of infection to ensure that individual cells contain only a single sgRNA, and cultures are grown for an extended period of time to allow for loss of cells with destruction of essential genes. The barcodes in the sgRNA constructs then allows their detection via DNA sequencing (*Fig. 6A*). sgRNAs against glyco-enzyme genes were compared to sgRNAs that destroy essential genes such as *MYC* as well as to non*-*relevant sgRNAs. As shown in *Fig. 6B*, compared to non*-*relevant sgRNAs (green circles), sgRNAs ablating many of the enzymes involved in glycan synthesis did not selectively disappear, indicating that these enzymes are not essential for cell survival in culture conditions. Other sgRNAs under-represented compared to the non*-*relevant sgRNAs included those targeting O*-*glycan synthesis (*GALNT2, GCNT1, C1GALT1, C1GALT1, Suppl. Table 6*). *MGAT1* ablation clearly reduced cell survival (*Fig. 6C*), and it ranked among the most critical genes, comparable to essential genes such as *MYC* (*Fig. 6D*). Interestingly, ablation of *OGT1* and *OGA*, enzymes responsible for application and removal of O*-*GlcNAc monosaccharides [60], and *NGLY1*, an enzyme located in the cytosol that removes N*-*glycans from glycoproteins subjected to proteasome degradation [61], was as lethal as that of essential genes such as MYC (*Fig. 6D*).

**Figure 6.**
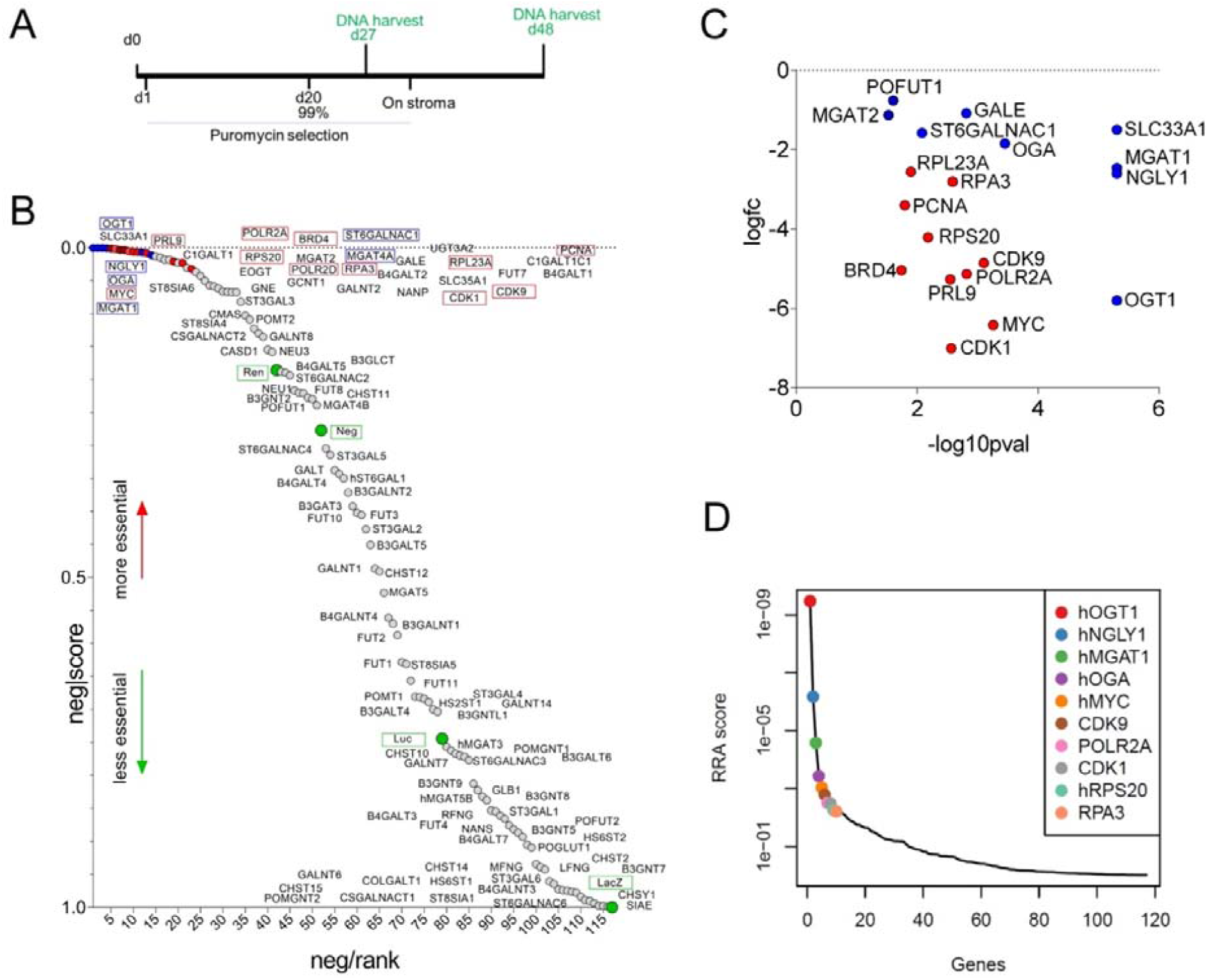
Cas9/CRISPR screen identifies enzymes involved in glycome remodelling essential for MLL-r B-ALL survival. (A) Schematic diagram of selective ablation of KOPN8 MLL-r BCP-ALL cells expressing Cas9 protein and sgRNAs for 102 glycan*-*building genes. (B) Negative-ranked MAGeCK score of genes involved in glycan building compared to genes with neutral selection activity (green circles, n=10 sgRNAs/gene) and genes that are essential for cell survival (red circles, n=2 sgRNAs/gene; names of genes indicated with a red outline). Labelling of points is approximate; for exact ranking see *Suppl. Table 6*. (C) Genes of which targeting sgRNAs were significantly (p<0.05) under-represented. (D). RRA score plot showing the overall combined ranking of top ten genes needed for survival of KOPN8 cells. BRD4, CDK1, CDK9, MYC, PCNA, POLR2A, POLR2D, PRL9, RPS20 and RPA3 are control genes included in the screen as essential for cell survival. LacZ, Luc, Neg and Ren sgRNAs are passive passengers in transduced cells and are not under selective negative pressure.

### Transcriptomic and proteomic data show typically close correlation

To date no in*-*depth proteomic screening has been reported for patient-derived human primary MLL-r cells. The current knowledge on protein changes in leukemic MLL-r compared to normal precursor B cells is largely based on very few reported transcriptomics studies in which matched normal controls were included as a benchmark to evaluate differences [3, 62, 63]. Therefore, we analysed the total MLL-r proteome and compared it to that of control cells to evaluate how well the transcriptome and proteome data correlate. Overall, 408 proteins exhibited significantly different levels in the label-free quantitation (LFQ) analyses (FDR≤1 % and log2FC≥2), of which 206 were upregulated in MLL-r cells (*Fig. 7A*). This included 40 proteins that were previously reported to be O*-*glycosylated in other cell types [42]. Unsurprisingly, a higher number of differentially-expressed transcripts were reported from the RNA-seq analyses and identified 3936 genes with differential expression at an mRNA level (FDR≤5 % and log2FC≥1), with 1883 genes upregulated in MLL-r cells (*Suppl. Table 1*). High pH-fractionation proteomics provided clear evidence that approximately 10 % differentially regulated genes were observed.

**Figure 7.**
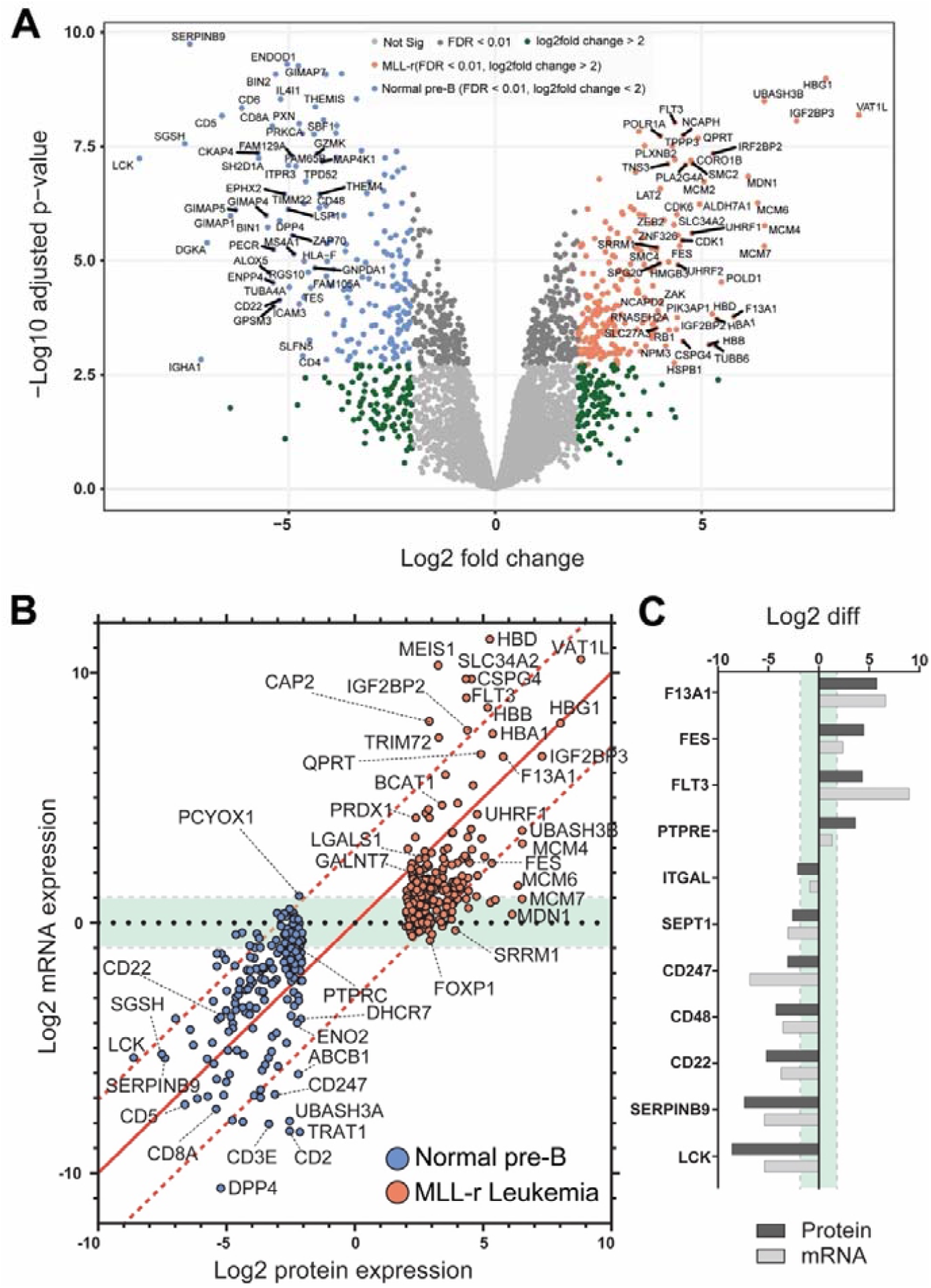
MLL-r and normal B-cell precursor controls show concordant proteome and transcriptome changes. (A) Volcano plot representing all 4225 proteins (with a median of 3202 proteins per sample; *Suppl. Table 2*) identified by orthogonal, high pH-fractionation. The top 50 most differentially expressed genes for each group are indicated. Proteins overexpressed in MLL, red; downregulated proteins, blue. (B) Correlation plot between protein and mRNA expression showing tight quantitative correlation (227 proteins; ≈57 %; log-fold difference <3) and good correlation (29 proteins; ≈7 %; similar up or down*-*regulation trend but >3-fold log fold difference). Green area: 84/141 remaining genes (≈36 %) with concordant differential expression but with very low mRNA levels (also see *Suppl. Table 7*). PCYOX1, the sole gene with negative correlation (proteomics: downregulated; RNA-seq: upregulated) between the two datasets. The 1:1 correlation axis is indicated with a red line. (C) Specific examples of correlated protein and mRNA levels.

To investigate the correlation between our transcriptomic and proteomic datasets we plotted Log2 value changes of RNA against protein changes of the 397 differentially-expressed proteins identified by both techniques (*Fig. 7B and 7C*). Out of these, a very good correlation between the expression of protein and transcript was found for 227 proteins (*Fig. 7B*).

Previous RNA expression analyses for BCP-ALL that included MLL-r samples and normal controls [3, 62, 63] reported increased expression of hallmark genes such as FLT3 [64] and CSPG4 [65] which was confirmed in our study both by transcriptomics and LFQ proteomics (*Fig. 7A* and *7B*). FLT3, for example, was almost 9 log-fold upregulated at the RNA-seq level, but just 4.35 log-fold upregulated in the proteomics dataset (*Fig. 7C*). Integrin α4 (*ITGA4*), for which increased expression has been previously associated with very poor outcomes in BCP-ALL [66], was the only integrin out of the ten detected that was upregulated in MLL-r cells at the transcript level. Transcriptomics data also showed an increased expression of mucin*-*like glycoproteins including CD43 (*SPN*) and decreased expression of P-selectin glycoprotein ligand-1 (*SELPLG*) (*Suppl. Table 2*). Both are highly-glycosylated proteins, and these modifications may have contributed to why they were not detected in our proteomics dataset. In fact, one CD43 peptide (363-400) was identified in the proteomics dataset but excluded for further consideration due to our minimum requirement of two or more unique peptides for positive protein identification.

## Discussion

Our studies clearly show that the glycocalyx is rich in hitherto unexplored diagnostic and therapeutic targets that remain elusive in gene/transcriptome-focused screening evaluations. Primary MLL-r cells that have a block in differentiation compared to normal precursor B-lineage controls, show a vastly increased O*-*glycome complexity. Major shifts in the MLL-r glycocalyx include a strong increase of Core 2 type O*-*glycans (*Fig. 2B*), as well as an overall increase in sialylated N*-* glycans (*Fig. 3B*). These changes were accompanied by several alterations of the cell glycosylation machinery, determined at both the gene expression and glycosyltransferase levels. Remarkably, the Core 1 to Core 2 shift in the O-glycome of MLL-r cells seems to be mainly a consequence of the expression of an important transferase, GCNT1 which was upregulated at the transcript level in MLL-r samples (*Fig. 1C*). These results show a striking similarity to the observations made in CML and AML cells, in which an increased activity and expression of GCNT1 led to an increase in Core 2 O*-*glycans, as compared to normal granulocytes [67]. Interestingly, Giovannone *et al* investigated protein O-glycosylation in mature peripheral normal and malignant B-cells and found that less differentiated B-cells also have higher O-glycan complexity correlating with higher GCNT1 expression [68], supporting the notion that differentiation of B-cells in general reduces the complexity of O-glycans.

Furthermore, we found that a specific glycosyltransferase, GALNT7, which is one of the O*-*GalNAc glycosylation*-*initiating enzymes, was significantly upregulated in patient samples at both transcript and protein levels (*Fig. 1A* and *1B*) as corroborated by gene expression array data (*Suppl. Fig. 4*). Although this enzyme has previously been linked to carcinogenesis in cultured cell models of colorectal and breast cancer [for example [69, 70]], this is the first time that GALNT7 was shown to be increased at both a transcript and protein level in primary patient MLL-r cells compared to normal controls. Yeoh *et al* [71] also reported a significant loss of *GALNT7* mRNA in conjunction with a reduced leukemia burden in B-cell precursor patient samples during the course of initial induction chemotherapy (also see *Suppl. Fig. 5*). GALNT7 (also known as ppGalNAcT7) was reported to have a strict substrate preference, and to glycosylate peptides not glycosylated by any other ppGalNAcT isoform [37]. This supports the notion that potential protein substrates such as CD45 [68], CD43 or CD44 in MLL-r cells could undergo site-specific O-glycosylation events that are absent in normal BCP cells and which could be used as a leukemia-specific marker or target for treatment.

We also identified increased levels of sialylated N-glycans in MLL-r cells. This is of interest given that that aberrant sialylation, with an emphasis on altered expression of ST6GAL1, has been extensively related to malignant transformation [as reviewed in [72, 73]]. Here we observed increased overall levels of sialylated N-glycans in MLL-r cells with increases in both to α2-6 and α2-3 linked sialic acid-linked glycoconjugates (*Fig. 3B*). Intriguingly, although MLL-r cells contained higher levels of α2-6 sialic acid containing N-glycans, ST6GAL1 transcript and protein levels were significantly lower in these patient cells, suggesting that lower ST6Gal1 levels are still sufficient to modify N-glycans with α2-6 linked NeuAc residues, or that alternative pathways are present in MLL-r cells that maintain the overall α2-6 NeuAc levels. The results of our CRISPR screen testing of the enzymes involved in glycan synthesis is consistent with the latter possibility. Interestingly, T-cell ALL cells made resistant to the chemotherapeutic agent desoxyepothilone B have decreased levels of α2-6 NeuAc residues on their membrane glycoproteins, which correlated with reduced ST6Gal1 activity and mRNA expression [74].

Fucosylation was also altered in MLL-r cells. Interestingly, CD15, a Lewis X antigen detected by flow cytometry, is regarded as a hallmark of MLL-r BCP-ALL [75]. We found that compared to normal BCP cells, levels of N-glycan associated Lewis X type fucose were significantly higher in the MLL-r samples, with FUT4, one of the enzymes responsible for Lewis X synthesis, being increased more than threefold (*Fig. 3E* and *3F*, also see *Suppl. Fig. 4*). Increased *FUT4* RNA also is included in the signature that clusters MLL-r leukemias together from other subclasses [3] and was suggested to regulate migration and adhesion of B-ALL cells by integrin α5β1 [76]. Thus, we believe that increased Lewis X synthesis could contribute to MLL-AF4-driven migration and invasion [77].

GAGs are extremely important biomolecules of the extracellular matrix, as they modulate protein function and stability. Tsidulko *et al* reported that many proteoglycans are expressed in EBV-transformed normal and malignant mature B-cell lines [78], and most studies investigating GAGs have similarly focused on *in vitro* cultured leukemia cell lines [79]. Makatsori *et al* showed that the distribution of CS and HS differed between various leukemic cell lines [80]. They also showed that CS was the more abundant GAG secreted in cell culture medium while the cellular levels of CS and HS were roughly comparable. GAGs have also been shown to be important for the ability of hematopoietic progenitor cells to bind to bone marrow endothelial cells [81], indicating an intrinsic relevance for GAGs in specific stages of immune cell maturation.

Our RNA-seq results suggest that MLL-r cells partly modulate their GAG biosynthesis pathways from CS to HS (*Fig. 5A*). Interestingly, of the six known CS proteoglycans, CSPG4 (NG2), which is diagnostic for the MLL-r subtype of B-cell precursor ALL [82, 83], is also strongly overexpressed in our proteomics dataset. CSPG4 has a single O*-*linked CS chain attached at S995, with an oncofetal composition in many cancer cell types and mediating adhesion to integrins α4, β1 and α5β1 [84]. CHST11 (CS-specific sulfotransferase 11) is required for the synthesis of this CS structure [85] and we found this enzyme to be overexpressed in MLL-r cells compared to normal controls (*Fig. 5A*). Thus, CSPG4 may play an important role in MLL-r leukemia cell adhesion. Indeed, a recent study using a PDX model of MLL-r ALL showed that enzymatic removal of the CS chain from CSPG4 enhanced the effects of standard chemotherapy in mice [86], suggesting the observed chemosensitisation by reduced integrin*-*mediated adhesion in the bone marrow microenvironment.

In agreement with previous reports [54, 55], we observed high levels of Galectin*-*1 in MLL-r samples (2.75-fold higher, *Fig. 4C*), as well as higher levels of Galectin*-*1 binding epitopes on these cells (*Fig. 4A, B*). It has been suggested that Galectin*-*1 can also interact with chondroitin sulfate B and might be involved in the extracellular matrix assembly in smooth muscle cells [87]. Given the strong upregulation of Galectin*-*1 in MLL-r cells, a cell surface remodelling towards higher levels of HS might also play an important role in MLL-r cell survival within the bone marrow environment.

Many of the heparan sulfate proteoglycans (HSPGs) are expressed at relatively high levels in normal and leukemic B-cell precursors (*Suppl. Table 2*). *SPOCK2* mRNA, encoding a secreted HSPG [88] was especially abundant, and *SDC2* and *HSPG2* were significantly higher in the MLL-r samples (*Suppl. Table 2)*. Interestingly, *HSPG2* was included in the group of 72 genes downregulated on d8 in a cohort of pediatric patients treated with chemotherapy and designated by Yeoh *et al* as predictive of overall response [71]. HSPGs are key regulators of the bone marrow niche of normal hematopoietic stem cells [89], and thus the proposed cell surface remodelling from CS to HS synthesis could regulate both extracellular protein modification as well as change the GAG composition of those proteins modified by both CS as well as HS such as TGFBR3 [88].

We also compared RNA expression data with protein expression in the same cells. Within the proteomics dataset, we reliably confirmed protein level changes for about 11 % of all transcripts identified to be differentially regulated in MLL-r cells. Within these 11 %, we also identified, and reliably quantified, proteins generally considered to be present at low abundance levels, such as Golgi-bound glycosyltransferases (*Fig. 2-4*). The overall trend with respect to up- or downregulation was in good agreement between the techniques for about 80 % of the detected proteins/transcripts. Nevertheless, proteomics delivered a different quantitative result for around 19 % of proteins and identified six proteins not detected by transcriptomics. Thus, there is clearly a significant value in applying both approaches as these deliver orthogonal information and cross-validate findings. Of the 11 proteins reliably identified and found to exhibit significant changes solely in the proteomics data, corresponding values were not present in the RNA-seq data. In part, this was due to the fact that the transcripts were not included in the UCSC annotation of hg38 (P0DTU4, a specific T-cell receptor beta chain) or were excluded from automated downstream analysis because they were not annotated as protein coding (IGHA1, IGHM). However, for others, RNA expression levels fell below the cut-off threshold of <1.0 (ACY1, CES1, EPX, ITGA2B, ITGB3, MMS22L, PRG2, VTN) indicating that high levels of specific proteins were present despite low to non*-*existent levels of the corresponding mRNA. We speculate that this could be due to recruitment of protein from exogenous sources within the bone marrow microenvironment. For example, vitronectin (VTN) and PRG2 are both well-known to bind to HS chains [90, 91], indirectly supporting our transcriptomics findings which suggests GAG-remodelling into a HS-rich glycocalyx in MLL-r cells. Similarly, eosinophil peroxidase (EPX), a protein rich in calcium and strongly expressed in bone marrow, could associate with strongly negatively-charged GAG-chains via charge interactions. ITGA2B and ITGB3 encode integrins that form the platelet glycoprotein (GP)IIb/IIIa [92], and their presence is most likely caused by platelet residues in the protein preparations. The lower integrin α2b and β3 (ITGA2B, ITGB3) protein levels found in the MLL-r samples (*Suppl. Table 7*) is consistent with the presumed BM microenvironment origin of these proteins: they would be expected to be present at lower levels in the BM of leukemia cases, which typically present with thrombocytopenia. Thus, our analyses of the primary leukemia samples may have detected changes in the tumour microenvironment that are invisible in cell-culture based experiments.

We found that the expression of many genes involved in glycosylation differ between MLL-r cells and normal controls: 61 of the 221 human glycosyltransferases are differentially expressed in MLL-r cells. Some of these differences are likely to be caused by the overall modified metabolic state of leukemic cells compared to the corresponding normal controls, but a subset could also be aberrantly transcribed due to the fusion of the *MLL* gene [also known as *KMT2a*] with different partners. Godfrey *et al* analysed potential target genes that contain an enhancer which can bind histone H3 with K29 methylation in among others the MLL-r B-ALL cell lines RS4;11 and SEM [93]. Such genes are candidates for deregulation by the recruitment of MLL1/KMT2A fusion proteins to the DNA through interaction with the histone H3K79 methyltransferase DOT1L. We found that 865 genes with differential expression between MLL-r and controls in our RNA-seq analyses contain such enhancers including genes known to have aberrant expression such as *CSPG4* and *MEIS1*. Interestingly, candidate MLL-DOT1L-regulated genes include those encoding enzymes involved in the O-glycosylation (GCNT1, GALNT1, GALNT2, GALNT7) and N-glycosylation (MGAT1, MGAT4A, MGAT 4B, and MGAT 5) pathways [not shown].

Our survey of survival-associated genes involved in glycan remodelling showed that those encoding enzymes that add terminal modifications such as sialylation and fucosylation have functional redundancy or are otherwise not critical for MLL-r cell survival under steady-state growth in culture. In contrast, functioning of the *MGAT1* gene encoding the first branching enzyme in the N-glycan biosynthesis pathway was clearly required for cell survival. This result was unexpected seeing that knockout of *mgat1* in mice is compatible with embryonic development until mid-gestation [94]. In addition, shRNA-mediated *MGAT1* knockdown in a human prostate cancer cell line was not lethal to these cells [95]. We were also able to identify three other genes of which the function is critical to MLL-r B-ALL cells. High levels of OGT protein expression were measured in controls and MLL-r samples (*Fig. 3D*) and significantly lower *OGA/MGEA5* RNA levels were found in the MLL-r samples (*Suppl. Table 2, Suppl. Fig. 7*). Homeostasis of O*-*GlcNAc through activities of OGT and OGA has been previously implicated in normal hematopoietic development [96, 97], in differentiation arrest of AML and Jurkat T-cells [98] as well as in other cancers [99]. However, our study is the first to report that this modification is also essential for the survival of MLL-r B-ALL cells. The essential function of NGLY1 for MLL-r cell survival under conditions of steady-state growth was unexpected because human patients with NGLY1 loss-of-function exist [100]. This enzyme is a non*-*redundant deglycosylase needed for the proteasome processing of misfolded proteins [101], with an essential function for AML cell survival [102]. Interestingly, Tomlin *et al* reported the development of a small molecule inhibitor of NGLY1 that enhanced cytotoxicity of the proteasome inhibitor bortezomib used to treat the mature B-cell malignancies multiple myeloma and mantle cell lymphoma [103]. Our results suggest such drug combination approaches may have specific lethality for MLL-r B-cell precursor ALLs as well.

In summary we describe the first multi-*omics* study, covering the transcriptome, proteome and glycome, from limited numbers (≈10^6^ cells per analysis) of primary MLL-r patient cells. While aspects of our study are consistent with previous observations concerning specific protein markers (FLT3, CSPG4, LGALS1) [64, 65], we have also found significantly different expression of other proteins such as the tyrosine kinase Fes. Interestingly, Kohlman *et al* reported that Fes would discriminate between AML and ALL with 11q rearrangement [104]. However, in our analysis we found that Fes protein and mRNA are higher in leukemic B-cell precursor cells compared to normal controls, suggesting Fes as a general target for MLL-rearranged leukemias. High expression of Fes in BCP-ALL with MLL-r rearrangement compared to normal BM controls is also consistent with published data (*Suppl. Fig 6)*. Thus, these results suggest that the development of a small molecule inhibitor targeting this kinase could be useful to treat MLL-rearranged leukemias. For example, a combination therapy with inhibitors of DOT1L or of the DOT1L/MENIN interaction, that are in clinical trials for the treatment of MLL-r acute leukemias [105], could be an exciting new therapeutic approach.

Aberrant protein glycosylation has been extensively linked to malignancy across every type of cancer known to date [reviewed in [12, 106, 107]]. Importantly, even minor alterations in the glycan structure can strongly change the activity of glycoconjugates and through this mechanism alter cell behaviour. Here we present the first combined, multi-*omics* study employing transcriptomics, proteomics and glycomics to capture the protein and glycan landscape of primary non*-*cultured MLL-r patient cells. This first comprehensive multi-*omics* study on MLL-r thus also represents a valuable, novel data resource. We find that the leukemia cell glycome, together with many proteins associated with glycan biosynthesis, are significantly changed compared to normal control precursor B-cells, indicating that the malignant phenotype of these cells could be regulated by such changes. We highlight the relevance of using primary, patient derived cells to identify leukemia-associated proteins invisible to gene-centric approaches, and demonstrate how integrated, multi-*omics* screening delivers a novel, inclusive map of the MLL-r cell landscape. Future studies therefore are warranted to determine how glycans on proteins have been remodelled and how these can be specifically targeted.

## Conclusions

This first multi*-omics* analysis of primary MLL-r BCP-ALL leukemia cells revealed a global reorganisation of the MLL-r cell glycocalyx. Using an integrated multi-*omics* workflow did not just enable us to discover previously unidentified diagnostic/therapeutic protein targets for MLL-r, but also revealed that a multi*-omics* approach delivers an added benefit to identify novel markers that are not detectable solely by a transcriptomics approach.

## Data Availability

Glycomics data can be access on GlycoPOST using the following credentials:
URL: https://glycopost.glycosmos.org/preview/192552055960853e1fc17f0
PIN code: 5775
Proteomics data can be accessed and reviewed on PRIDE using the following credentials:
URL: https://www.ebi.ac.uk/pride
Project Name: Altered glycoproteome of MLL-rearranged B-cell precursor acute lymphoblastic leukemia
The Transcriptomics data has been deposited in GEO, accession Nr. GSE176364:
Go to https://www.ncbi.nlm.nih.gov/geo/query/acc.cgi?acc=GSE176364, Enter token mnidkuosnzavxer
Project accession: PXD025354
Username:
Password: FldtRJ8T

https://www.ebi.ac.uk/pride

https://www.ncbi.nlm.nih.gov/geo/query/acc.cgi?acc=GSE176364

## Declarations

### Ethics approval and consent to participate

All human specimen collection protocols were reviewed by Children’s Hospital Los Angeles Institution Review Board (IRB) and Committee on Clinical Investigations (CCI). Protocols for collection were approved and found to be in compliance with ethical practices by the Children’s Hospital Los Angeles Institution Review Board and Committee on Clinical Investigations. All specimens were de-identified/anonymized before acquisition for research. The specimens were collected as leftover specimens that were initially collected for clinical diagnostic purposes and were discarded as medical waste when no longer needed for clinical purposes.

### Availability of data and materials

Preview of the glycan data can be access on GlycoPOST(11) using the following credentials: URL:https://glycopost.glycosmos.org/preview/192552055960853e1fc17f0

PIN CODE: 5775

The glycomics data has also been incorporated into GlyConnect https://beta.glyconnect.expasy.org/browser/references/2975, a platform integrating sources of information to help characterise the molecular components of protein glycosylation [108].

The mass spectrometry proteomics data can be accessed and reviewed on PRIDE using the following credentials

Username

Password: FldtRJ8T

The Transcriptomics data has been deposited in GEO, accession Nr. GSE176364:

Go to https://www.ncbi.nlm.nih.gov/geo/query/acc.cgi?acc=GSE176364, Enter token mnidkuosnzavxer

All transcriptomics, proteomics and glycomics data results are also provided as supplementary material.

## Acknowledgements

We thank Sachith Gallolu for analysis of the GSE67684 data. The SC2 Core and Li Fan at Children’s Hospital Los Angeles is acknowledged for RNA-seq analysis. We also would like to thank Frédérique Lisacek and Catherine Hayes from the Swiss Institute of Bioinformatics, Glycomics@Expasy portal, for their support in integrating the glycomics data into GlyConnect.

The authors declare no competing interests.

## Conceptualization

TO, NH and DK; Glycomics and Proteomics: TO, KA, AA, NP, DK; RNA-Seq: EJJ; Primary Cells: HAA, EJJ, CHC; CRISPR experiments: MZ, CWC, LY; JC, XQ. Formal Analysis and Investigation: TO, NH, DK; Bioinformatics: TO, FJ, LY, NH; Manuscript conceptualisation and writing: TO, NH, DK; All authors reviewed the manuscript and provided comments. Funding Acquisition: MvI, NH, DK; Supervision: NH, DK

## References

1. Sung H, Ferlay J, Siegel RL, Laversanne M, Soerjomataram I, Jemal A, et al. Global Cancer Statistics 2020: GLOBOCAN Estimates of Incidence and Mortality Worldwide for 36 Cancers in 185 Countries. CA: a cancer journal for clinicians. 2021; 71: 209–49.

2. Pui CH, Robison LL, Look AT. Acute lymphoblastic leukaemia. Lancet. 2008; 371: 1030–43.

3. Gu Z, Churchman ML, Roberts KG, Moore I, Zhou X, Nakitandwe J, et al. PAX5-driven subtypes of B-progenitor acute lymphoblastic leukemia. Nature genetics. 2019; 51: 296–307.

4. Meyer C, Burmeister T, Groger D, Tsaur G, Fechina L, Renneville A, et al. The MLL recombinome of acute leukemias in 2017. Leukemia. 2018; 32: 273–84.

5. Pui CH. Precision medicine in acute lymphoblastic leukemia. Front Med. 2020; 14: 689–700.

6. Hilden JM, Dinndorf PA, Meerbaum SO, Sather H, Villaluna D, Heerema NA, et al. Analysis of prognostic factors of acute lymphoblastic leukemia in infants: report on CCG 1953 from the Children’s Oncology Group. Blood. 2006; 108: 441–51.

7. Rayes A, McMasters RL, O’Brien MM. Lineage Switch in MLL-Rearranged Infant Leukemia Following CD19-Directed Therapy. Pediatr Blood Cancer. 2016; 63: 1113–5.

8. Mullighan CG. Molecular genetics of B-precursor acute lymphoblastic leukemia. J Clin Invest. 2012; 122: 3407–15.

9. Steinhilber D, Marschalek R. How to effectively treat acute leukemia patients bearing MLL-rearrangements ? Biochem Pharmacol. 2018; 147: 183–90.

10. Verma D, Zanetti C, Godavarthy PS, Kumar R, Minciacchi VR, Pfeiffer J, et al. Bone marrow niche-derived extracellular matrix-degrading enzymes influence the progression of B-cell acute lymphoblastic leukemia. Leukemia. 2020; 34: 1540–52.

11. Chin-Hun Kuo J, Gandhi JG, Zia RN, Paszek MJ. Physical biology of the cancer cell glycocalyx. Nat Phys. 2018; 14: 658–69.

12. Pinho SS, Reis CA. Glycosylation in cancer: mechanisms and clinical implications. Nature reviews Cancer. 2015; 15: 540–55.

13. Varki A, Kannagi R, Toole B, Stanley P. Glycosylation Changes in Cancer. In: rd, Varki A, Cummings RD, Esko JD, Stanley P, Hart GW, et al., editors. Essentials of Glycobiology. 3rd ed. Cold Spring Harbor (NY): Cold Spring Harbor (NY); 2015. p. 597-609.

14. Cerliani JP, Blidner AG, Toscano MA, Croci DO, Rabinovich GA. Translating the ‘Sugar Code’ into Immune and Vascular Signaling Programs. Trends Biochem Sci. 2017; 42: 255–73.

15. Hobbs SJ, Nolz JC. Regulation of T Cell Trafficking by Enzymatic Synthesis of O-Glycans. Frontiers in immunology. 2017; 8: 600.

16. Reily C, Stewart TJ, Renfrow MB, Novak J. Glycosylation in health and disease. Nat Rev Nephrol. 2019; 15: 346–66.

17. Haferlach T, Kohlmann A, Wieczorek L, Basso G, Kronnie GT, Bene MC, et al. Clinical utility of microarray-based gene expression profiling in the diagnosis and subclassification of leukemia: report from the International Microarray Innovations in Leukemia Study Group. J Clin Oncol. 2010; 28: 2529–37.

18. Bagger FO, Sasivarevic D, Sohi SH, Laursen LG, Pundhir S, Sonderby CK, et al. BloodSpot: a database of gene expression profiles and transcriptional programs for healthy and malignant haematopoiesis. Nucleic acids research. 2016; 44: D917–24.

19. Wessel D, Flugge UI. A method for the quantitative recovery of protein in dilute solution in the presence of detergents and lipids. Anal Biochem. 1984; 138: 141–3.

20. Jensen PH, Karlsson NG, Kolarich D, Packer NH. Structural analysis of N-and O-glycans released from glycoproteins. Nat Protoc. 2012; 7: 1299–310.

21. Everest-Dass AV, Abrahams JL, Kolarich D, Packer NH, Campbell MP. Structural feature ions for distinguishing N-and O-linked glycan isomers by LC-ESI-IT MS/MS. J Am Soc Mass Spectrom. 2013; 24: 895–906.

22. Hinneburg H, Korac P, Schirmeister F, Gasparov S, Seeberger PH, Zoldos V, et al. Unlocking Cancer Glycomes from Histopathological Formalin-fixed and Paraffin-embedded (FFPE) Tissue Microdissections. Mol Cell Proteomics. 2017; 16: 524–36.

23. Sinha D, Mandal C, Bhattacharya DK. Identification of 9-O acetyl sialoglycoconjugates (9-OAcSGs) as biomarkers in childhood acute lymphoblastic leukemia using a lectin, AchatininH, as a probe. Leukemia. 1999; 13: 119–25.

24. Parameswaran R, Lim M, Arutyunyan A, Abdel-Azim H, Hurtz C, Lau K, et al. O-acetylated N-acetylneuraminic acid as a novel target for therapy in human pre-B acute lymphoblastic leukemia. J Exp Med. 2013; 210: 805–19.

25. Kolarich D, Rapp E, Struwe WB, Haslam SM, Zaia J, McBride R, et al. The minimum information required for a glycomics experiment (MIRAGE) project: improving the standards for reporting mass-spectrometry-based glycoanalytic data. Mol Cell Proteomics. 2013; 12: 991–5.

26. York WS, Agravat S, Aoki-Kinoshita KF, McBride R, Campbell MP, Costello CE, et al. MIRAGE: the minimum information required for a glycomics experiment. Glycobiology. 2014; 24: 402–6.

27. Struwe WB, Agravat S, Aoki-Kinoshita KF, Campbell MP, Costello CE, Dell A, et al. The minimum information required for a glycomics experiment (MIRAGE) project: sample preparation guidelines for reliable reporting of glycomics datasets. Glycobiology. 2016; 26: 907–10.

28. Campbell MP, Abrahams JL, Rapp E, Struwe WB, Costello CE, Novotny M, et al. The minimum information required for a glycomics experiment (MIRAGE) project: LC guidelines. Glycobiology. 2019; 29: 349–54.

29. Varki A, Cummings RD, Aebi M, Packer NH, Seeberger PH, Esko JD, et al. Symbol Nomenclature for Graphical Representations of Glycans. Glycobiology. 2015; 25: 1323–4.

30. Neelamegham S, Aoki-Kinoshita K, Bolton E, Frank M, Lisacek F, Lutteke T, et al. Updates to the Symbol Nomenclature for Glycans guidelines. Glycobiology. 2019; 29: 620–4.

31. Sanjana NE, Shalem O, Zhang F. Improved vectors and genome-wide libraries for CRISPR screening. Nat Methods. 2014; 11: 783–4.

32. Uckelmann HJ, Kim SM, Wong EM, Hatton C, Giovinazzo H, Gadrey JY, et al. Therapeutic targeting of preleukemia cells in a mouse model of NPM1 mutant acute myeloid leukemia. Science. 2020; 367: 586–90.

33. Li W, Xu H, Xiao T, Cong L, Love MI, Zhang F, et al. MAGeCK enables robust identification of essential genes from genome-scale CRISPR/Cas9 knockout screens. Genome biology. 2014; 15: 554.

34. Tyanova S, Temu T, Cox J. The MaxQuant computational platform for mass spectrometry-based shotgun proteomics. Nat Protoc. 2016; 11: 2301–19.

35. Shah AD, Goode RJA, Huang C, Powell DR, Schittenhelm RB. LFQ-Analyst: An Easy-To-Use Interactive Web Platform To Analyze and Visualize Label-Free Proteomics Data Preprocessed with MaxQuant. J Proteome Res. 2020; 19: 204–11.

36. Bennett EP, Mandel U, Clausen H, Gerken TA, Fritz TA, Tabak LA. Control of mucin-type O-glycosylation: a classification of the polypeptide GalNAc-transferase gene family. Glycobiology. 2012; 22: 736–56.

37. Kong Y, Joshi HJ, Schjoldager KT, Madsen TD, Gerken TA, Vester-Christensen MB, et al. Probing polypeptide GalNAc-transferase isoform substrate specificities by in vitro analysis. Glycobiology. 2015; 25: 55–65.

38. Steentoft C, Vakhrushev SY, Joshi HJ, Kong Y, Vester-Christensen MB, Schjoldager KT, et al. Precision mapping of the human O-GalNAc glycoproteome through SimpleCell technology. The EMBO journal. 2013; 32: 1478–88.

39. Ju T, Cummings RD. A unique molecular chaperone Cosmc required for activity of the mammalian core 1 beta 3-galactosyltransferase. Proceedings of the National Academy of Sciences of the United States of America. 2002; 99: 16613–8.

40. Ju T, Brewer K, D’Souza A, Cummings RD, Canfield WM. Cloning and expression of human core 1 beta1,3-galactosyltransferase. J Biol Chem. 2002; 277: 178–86.

41. Autelitano F, Loyaux D, Roudieres S, Deon C, Guette F, Fabre P, et al. Identification of novel tumor-associated cell surface sialoglycoproteins in human glioblastoma tumors using quantitative proteomics. PloS one. 2014; 9: e110316.

42. Yang W, Ao M, Hu Y, Li QK, Zhang H. Mapping the O-glycoproteome using site-specific extraction of O-linked glycopeptides (EXoO). Molecular systems biology. 2018; 14: e8486.

43. Kudelka MR, Nairn AV, Sardar MY, Sun X, Chaikof EL, Ju T, et al. Isotopic labeling with cellular O-glycome reporter/amplification (ICORA) for comparative O-glycomics of cultured cells. Glycobiology. 2018; 28: 214–22.

44. Stanley P, Taniguchi N, Aebi M. N-Glycans. In: rd, Varki A, Cummings RD, Esko JD, Stanley P, Hart GW, et al., editors. Essentials of Glycobiology. Cold Spring Harbor (NY): Cold Spring Harbor Laboratory Press; 2015. p. 99–111.

45. Alatrash G, Qiao N, Zhang M, Zope M, Perakis AA, Sukhumalchandra P, et al. Fucosylation Enhances the Efficacy of Adoptively Transferred Antigen-Specific Cytotoxic T Lymphocytes. Clin Cancer Res. 2019; 25: 2610–20.

46. Almeida A, Kolarich D. The promise of protein glycosylation for personalised medicine. Biochimica et biophysica acta. 2016; 1860: 1583–95.

47. Taniguchi N. HK, Fukuda M., Narimatsu H., Yamaguchi Y., Angata T. (eds.) 2nd edition. Handbook of Glycosyltransferases and Related Genes; 2014.

48. Mondal N, Dykstra B, Lee J, Ashline DJ, Reinhold VN, Rossi DJ, et al. Distinct human alpha(1,3)-fucosyltransferases drive Lewis-X/sialyl Lewis-X assembly in human cells. J Biol Chem. 2018; 293: 7300–14.

49. Gao C, Hanes MS, Byrd-Leotis LA, Wei M, Jia N, Kardish RJ, et al. Unique Binding Specificities of Proteins toward Isomeric Asparagine-Linked Glycans. Cell Chem Biol. 2019; 26: 535–47 e4.

50. Yang RY, Rabinovich GA, Liu FT. Galectins: structure, function and therapeutic potential. Expert Rev Mol Med. 2008; 10: e17.

51. Lichtenstein RG, Rabinovich GA. Glycobiology of cell death: when glycans and lectins govern cell fate. Cell Death Differ. 2013; 20: 976–86.

52. Giovannone N, Smith LK, Treanor B, Dimitroff CJ. Galectin-Glycan Interactions as Regulators of B Cell Immunity. Frontiers in immunology. 2018; 9: 2839.

53. Ruvolo PP. Galectins as regulators of cell survival in the leukemia niche. Adv Biol Regul. 2019; 71: 41–54.

54. Juszczynski P, Rodig SJ, Ouyang J, O’Donnell E, Takeyama K, Mlynarski W, et al. MLL-rearranged B lymphoblastic leukemias selectively express the immunoregulatory carbohydrate-binding protein galectin-1. Clin Cancer Res. 2010; 16: 2122–30.

55. Paz H, Joo EJ, Chou CH, Fei F, Mayo KH, Abdel-Azim H, et al. Treatment of B-cell precursor acute lymphoblastic leukemia with the Galectin-1 inhibitor PTX008. J Exp Clin Cancer Res. 2018; 37: 67.

56. Lin TW, Chang HT, Chen CH, Chen CH, Lin SW, Hsu TL, et al. Galectin-3 Binding Protein and Galectin-1 Interaction in Breast Cancer Cell Aggregation and Metastasis. J Am Chem Soc. 2015; 137: 9685–93.

57. Chen YH, Narimatsu Y, Clausen TM, Gomes C, Karlsson R, Steentoft C, et al. The GAGOme: a cell-based library of displayed glycosaminoglycans. Nat Methods. 2018; 15: 881–8.

58. Temkin V, Aingorn H, Puxeddu I, Goldshmidt O, Zcharia E, Gleich GJ, et al. Eosinophil major basic protein: first identified natural heparanase-inhibiting protein. The Journal of allergy and clinical immunology. 2004; 113: 703–9.

59. Overgaard MT, Haaning J, Boldt HB, Olsen IM, Laursen LS, Christiansen M, et al. Expression of recombinant human pregnancy-associated plasma protein-A and identification of the proform of eosinophil major basic protein as its physiological inhibitor. J Biol Chem. 2000; 275: 31128–33.

60. Hart GW. Three Decades of Research on O-GlcNAcylation - A Major Nutrient Sensor That Regulates Signaling, Transcription and Cellular Metabolism. Front Endocrinol (Lausanne). 2014; 5: 183.

61. Suzuki T, Huang C, Fujihira H. The cytoplasmic peptide:N-glycanase (NGLY1) - Structure, expression and cellular functions. Gene. 2016; 577: 1–7.

62. Hirabayashi S, Ohki K, Nakabayashi K, Ichikawa H, Momozawa Y, Okamura K, et al. ZNF384-related fusion genes define a subgroup of childhood B-cell precursor acute lymphoblastic leukemia with a characteristic immunotype. Haematologica. 2017; 102: 118–29.

63. Coustan-Smith E, Song G, Clark C, Key L, Liu P, Mehrpooya M, et al. New markers for minimal residual disease detection in acute lymphoblastic leukemia. Blood. 2011; 117: 6267–76.

64. Stubbs MC, Kim YM, Krivtsov AV, Wright RD, Feng Z, Agarwal J, et al. MLL-AF9 and FLT3 cooperation in acute myelogenous leukemia: development of a model for rapid therapeutic assessment. Leukemia. 2008; 22: 66–77.

65. Zangrando A, Intini F, te Kronnie G, Basso G. Validation of NG2 antigen in identifying BP-ALL patients with MLL rearrangements using qualitative and quantitative flow cytometry: a prospective study. Leukemia. 2008; 22: 858–61.

66. Hsieh YT, Gang EJ, Geng H, Park E, Huantes S, Chudziak D, et al. Integrin alpha4 blockade sensitizes drug resistant pre-B acute lymphoblastic leukemia to chemotherapy. Blood. 2013; 121: 1814–8.

67. Brockhausen I, Kuhns W, Schachter H, Matta KL, Sutherland DR, Baker MA. Biosynthesis of O-glycans in leukocytes from normal donors and from patients with leukemia: increase in O-glycan core 2 UDP-GlcNAc:Gal beta 3 GalNAc alpha-R (GlcNAc to GalNAc) beta(1-6)-N-acetylglucosaminyltransferase in leukemic cells. Cancer Res. 1991; 51: 1257–63.

68. Giovannone N, Antonopoulos A, Liang J, Geddes Sweeney J, Kudelka MR, King SL, et al. Human B Cell Differentiation Is Characterized by Progressive Remodeling of O-Linked Glycans. Frontiers in immunology. 2018; 9: 2857.

69. Mockl L, Pedram K, Roy AR, Krishnan V, Gustavsson AK, Dorigo O, et al. Quantitative Super-Resolution Microscopy of the Mammalian Glycocalyx. Dev Cell. 2019; 50: 57–72 e6.

70. Sahasrabudhe NM, Lenos K, van der Horst JC, Rodriguez E, van Vliet SJ. Oncogenic BRAFV600E drives expression of MGL ligands in the colorectal cancer cell line HT29 through N-acetylgalactosamine-transferase 3. Biol Chem. 2018; 399: 649–59.

71. Yeoh AE, Li Z, Dong D, Lu Y, Jiang N, Trka J, et al. Effective Response Metric: a novel tool to predict relapse in childhood acute lymphoblastic leukaemia using time-series gene expression profiling. Br J Haematol. 2018; 181: 653–63.

72. Munkley J, Scott E. Targeting Aberrant Sialylation to Treat Cancer. Medicines (Basel). 2019; 6.

73. Zhang Z, Wuhrer M, Holst S. Serum sialylation changes in cancer. Glycoconj J. 2018; 35: 139–60.

74. Nakano M, Saldanha R, Gobel A, Kavallaris M, Packer NH. Identification of glycan structure alterations on cell membrane proteins in desoxyepothilone B resistant leukemia cells. Mol Cell Proteomics. 2011; 10: M111 009001.

75. Pui CH, Rubnitz JE, Hancock ML, Downing JR, Raimondi SC, Rivera GK, et al. Reappraisal of the clinical and biologic significance of myeloid-associated antigen expression in childhood acute lymphoblastic leukemia. J Clin Oncol. 1998; 16: 3768–73.

76. Yi L, Hu Q, Zhou J, Liu Z, Li H. Alternative splicing of Ikaros regulates the FUT4/Le(X)-alpha5beta1 integrin-FAK axis in acute lymphoblastic leukemia. Biochem Biophys Res Commun. 2019; 510: 128–34.

77. Stavropoulou V, Kaspar S, Brault L, Sanders MA, Juge S, Morettini S, et al. MLL-AF9 Expression in Hematopoietic Stem Cells Drives a Highly Invasive AML Expressing EMT-Related Genes Linked to Poor Outcome. Cancer Cell. 2016; 30: 43–58.

78. Tsidulko AY, Matskova L, Astakhova LA, Ernberg I, Grigorieva EV. Proteoglycan expression correlates with the phenotype of malignant and non-malignant EBV-positive B-cell lines. Oncotarget. 2015; 6: 43529–39.

79. Maurer AM, Han ZC, Dhermy D, Briere J. Glycosaminoglycans enhance human leukemic cell growth in vitro. Leuk Res. 1994; 18: 837–42.

80. Makatsori E, Karamanos NK, Papadogiannakis N, Hjerpe A, Anastassiou ED, Tsegenidis T. Synthesis and distribution of glycosaminoglycans in human leukemic B-and T-cells and monocytes studied using specific enzymic treatments and high-performance liquid chromatography. Biomedical chromatography : BMC. 2001; 15: 413–7.

81. Netelenbos T, van den Born J, Kessler FL, Zweegman S, Merle PA, van Oostveen JW, et al. Proteoglycans on bone marrow endothelial cells bind and present SDF-1 towards hematopoietic progenitor cells. Leukemia. 2003; 17: 175–84.

82. Harrer DC, Schuler G, Dorrie J, Schaft N. CSPG4-Specific CAR T Cells for High-Risk Childhood B Cell Precursor Leukemia. International journal of molecular sciences. 2019; 20.

83. Prieto C, Lopez-Millan B, Roca-Ho H, Stam RW, Romero-Moya D, Rodriguez-Baena FJ, et al. NG2 antigen is involved in leukemia invasiveness and central nervous system infiltration in MLL-rearranged infant B-ALL. Leukemia. 2018; 32: 633–44.

84. Clausen TM, Pereira MA, Al Nakouzi N, Oo HZ, Agerbaek MO, Lee S, et al. Oncofetal Chondroitin Sulfate Glycosaminoglycans Are Key Players in Integrin Signaling and Tumor Cell Motility. Mol Cancer Res. 2016; 14: 1288–99.

85. Salanti A, Clausen TM, Agerbaek MO, Al Nakouzi N, Dahlback M, Oo HZ, et al. Targeting Human Cancer by a Glycosaminoglycan Binding Malaria Protein. Cancer Cell. 2015; 28: 500–14.

86. Lopez-Millan B, Sanchez-Martinez D, Roca-Ho H, Gutierrez-Aguera F, Molina O, Diaz de la Guardia R, et al. NG2 antigen is a therapeutic target for MLL-rearranged B-cell acute lymphoblastic leukemia. Leukemia. 2019; 33: 1557–69.

87. Moiseeva EP, Williams B, Samani NJ. Galectin 1 inhibits incorporation of vitronectin and chondroitin sulfate B into the extracellular matrix of human vascular smooth muscle cells. Biochimica et biophysica acta. 2003; 1619: 125–32.

88. Iozzo RV, Schaefer L. Proteoglycan form and function: A comprehensive nomenclature of proteoglycans. Matrix Biol. 2015; 42: 11–55.

89. Papy-Garcia D, Albanese P. Heparan sulfate proteoglycans as key regulators of the mesenchymal niche of hematopoietic stem cells. Glycoconj J. 2017; 34: 377–91.

90. Lane DA, Flynn AM, Pejler G, Lindahl U, Choay J, Preissner K. Structural requirements for the neutralization of heparin-like saccharides by complement S protein/vitronectin. J Biol Chem. 1987; 262: 16343–8.

91. Nunes QM, Su D, Brownridge PJ, Simpson DM, Sun C, Li Y, et al. The heparin-binding proteome in normal pancreas and murine experimental acute pancreatitis. PloS one. 2019; 14: e0217633.

92. Botero JP, Lee K, Branchford BR, Bray PF, Freson K, Lambert MP, et al. Glanzmann thrombasthenia: genetic basis and clinical correlates. Haematologica. 2020; 105: 888–94.

93. Godfrey L, Crump NT, Thorne R, Lau IJ, Repapi E, Dimou D, et al. DOT1L inhibition reveals a distinct subset of enhancers dependent on H3K79 methylation. Nat Commun. 2019; 10: 2803.

94. Schachter H. Mgat1-dependent N-glycans are essential for the normal development of both vertebrate and invertebrate metazoans. Semin Cell Dev Biol. 2010; 21: 609–15.

95. Beheshti Zavareh R, Sukhai MA, Hurren R, Gronda M, Wang X, Simpson CD, et al. Suppression of cancer progression by MGAT1 shRNA knockdown. PloS one. 2012; 7: e43721.

96. Zhang Z, Parker MP, Graw S, Novikova LV, Fedosyuk H, Fontes JD, et al. O-GlcNAc homeostasis contributes to cell fate decisions during hematopoiesis. J Biol Chem. 2019; 294: 1363–79.

97. Abramowitz LK, Hanover JA. T cell development and the physiological role of O-GlcNAc. FEBS Lett. 2018; 592: 3943–9.

98. Kampa-Schittenhelm KM, Haverkamp T, Bonin M, Tsintari V, Buhring HJ, Haeusser L, et al. Epigenetic activation of O-linked beta-N-acetylglucosamine transferase overrides the differentiation blockage in acute leukemia. EBioMedicine. 2020; 54: 102678.

99. Itkonen HM, Loda M, Mills IG. O-GlcNAc Transferase - An Auxiliary Factor or a Full-blown Oncogene? Mol Cancer Res. 2021; 19: 555–64.

100. Owings KG, Lowry JB, Bi Y, Might M, Chow CY. Transcriptome and functional analysis in a Drosophila model of NGLY1 deficiency provides insight into therapeutic approaches. Human molecular genetics. 2018; 27: 1055–66.

101. Park H, Suzuki T, Lennarz WJ. Identification of proteins that interact with mammalian peptide:N-glycanase and implicate this hydrolase in the proteasome-dependent pathway for protein degradation. Proceedings of the National Academy of Sciences of the United States of America. 2001; 98: 11163–8.

102. Wang T, Yu H, Hughes NW, Liu B, Kendirli A, Klein K, et al. Gene Essentiality Profiling Reveals Gene Networks and Synthetic Lethal Interactions with Oncogenic Ras. Cell. 2017; 168: 890–903 e15.

103. Tomlin FM, Gerling-Driessen UIM, Liu YC, Flynn RA, Vangala JR, Lentz CS, et al. Inhibition of NGLY1 Inactivates the Transcription Factor Nrf1 and Potentiates Proteasome Inhibitor Cytotoxicity. ACS Cent Sci. 2017; 3: 1143–55.

104. Kohlmann A, Schoch C, Dugas M, Schnittger S, Hiddemann W, Kern W, et al. New insights into MLL gene rearranged acute leukemias using gene expression profiling: shared pathways, lineage commitment, and partner genes. Leukemia. 2005; 19: 953–64.

105. Perner F, Armstrong SA. Targeting Chromatin Complexes in Myeloid Malignancies and Beyond: From Basic Mechanisms to Clinical Innovation. Cells. 2020; 9.

106. Stowell SR, Ju T, Cummings RD. Protein glycosylation in cancer. Annual review of pathology. 2015; 10: 473–510.

107. Munkley J, Elliott DJ. Hallmarks of glycosylation in cancer. Oncotarget. 2016; 7: 35478–89.

108. Alocci D, Mariethoz J, Gastaldello A, Gasteiger E, Karlsson NG, Kolarich D, et al. GlyConnect: Glycoproteomics Goes Visual, Interactive, and Analytical. J Proteome Res. 2019; 18: 664–77.

