## Supplementary material for "Glycoproteome remodelling in MLL-rearranged B-cell precursor acute lymphoblastic leukemia": Sup Material (from another version of this preprint: version 1, 2021-06-30)

Multi*-omics* of MLL-rearranged B-cell precursor acute lymphoblastic leukemia reveals broad glycoproteome remodelling

**This PDF file includes:**

Supplementary Material and Methods

References (1 to 10)

Figs. S1 to S7

Tables S1 to S7

**Supplementary Material and Methods**

N- and O-glycan release for glycomics analyses

50 µg of (glyco)proteins were reduced by adding a volume of 500 mM of Dithiothreitol to each sample, to a final concentration of 20 mM, at 50°C for 1 hour. After cooling, samples were then subjected to alkylation using 40 mM Iodoacetamide in the dark at room temperature. The reaction was quenched by adding another 20 mM of DTT and incubating for 5 minutes.

Proteins were precipitated using the Chloroform:Methanol:Water separation as described previously(1). The resulting protein pellet was left to air dry for 10 minutes. 5 µL of 8 M urea were added to each sample to resuspend the protein pellet using intensive vortexing. The final concentration of urea was adjusted to 4 M by adding 5 µL of pure water (MQ‑H_2_O).

The urea dissolved glycoproteins were immobilised on a PVDF membrane (Immobilon*-*P, 0.44 µm pore, Merck Millipore) before N-glycans were released enzymatically using PNGase F (50 U in MQ-H_2_O) and incubating overnight at 37°C, as described previously(2).. The released N-glycans (60 µL) were removed and transferred to another tube before incubating the solution with 10 µL 100 mM ammonium acetate for 1 hour at room temperature, followed by immediate drying of the samples in a vacuum centrifuge (Thermo Savant Speedvac). N*-*glycans were reduced by adding 20 µL of 1 M sodium borohydride in 50 mM potassium hydroxide and incubating at 50°C for 3 h. The reduction reaction was stopped by adding 2 µL of glacial acetic acid (Sigma, Cat#A6283). The reduced N*-*glycans were then desalted by cation exchange chromatography (AG50W-X8 catio*N-*exchange resin, BioRad, Cat#142-1431) packed onto C18 ZipTips (Merck Millipore, Cat#ZTC18S960). The flowthrough containing the reduced and purified N*-*glycans was then dried in a vacuum centrifuge. 100 µL of high-grade methanol (Sigma, Cat#34860) was added to remove the residual borate salts, and the purified *N-*glycans were dried under vacuum. This step was repeated twice to ensure complete removal of borates.

After removal of N-glycans, O-glycans were released chemically by reductive β‑elimination from the same PVDF membrane spots. 20 µL of 0.5 M sodium borohydride in 50 mM potassium hydroxide were added to the methanol-prewetted membrane spots and the samples were incubated at 50°C for 18 hours. The reaction was stopped using 2 µL of glacial acetic acid and the released and reduced O-glycans were desalted as described for the N*-*glycans(2).

Before mass spectrometry analyses, N*-* and O-glycans were carbon cleaned off‑line using porous-graphitised carbon (PGC) material packed on top of C18 ZipTips to avoid any potential contaminants and then stored at -20°C until their PGC-LC-ESI-MS/MS analyses.

PGC-nanoLC-ESI-MS/MS glycomics

All glycomics analyses were performed on an amaZon speed Ion Trap Mass Spectrometer (IT-MS, Bruker) attached to an UltiMate™ 3000 nanoUHPLC (Thermo Scientific™). The amaZon IT-MS was equipped with a CaptiveSpray ion Source (Bruker, Bremen, Germany). The LC-system consisted of a Hypercarb™ HPLC precolumn (5 μm, 30 mm x 0.32 mm, Thermo Fisher Scientific, Cat#11504070) and separation Hypercarb™ nano column (3 μm, 100 mm x 0.075 mm, Thermo Fisher Scientific, Cat#11504070) that were both maintained at 45°C. 10 mM ammonium bicarbonate (solvent A) and 70% ACN in 10 mM ammonium bicarbonate (solvent B) were used as the mobile phases for glycan separation. The flow rate used for loading was 6 μL/min. Valve switch was programmed to 6.5 min. The flow rate used for the separation of both N- and O-glycans was 3 μL/min and 1.5 μL/min, respectively.

N-glycans were separated using the following gradient: 0-14 min, 1% B; 14-15 min, linear increase to 1.1% B; 15-16 min, linear increase to 15.8% B; 16-67.25 min, linear increase to 40.2% B. Washing and re-equilibration steps were performed: 67.25-70 min, linear increase to 90% B; 70-75 min, held at 90% B; 75-76 min, return to 1% B; 76-90 min, re-equilibration at 1% B. The second valve switch was programmed to 75 min. For *N-*glycans analysis in IT-MS, MS spectra were obtained in negative ion mode between a mass range of *m/z* 500-1800, using the instruments UltraScan mode. The smart parameter setting (SPS) defined target mass used was *m/z* 1250, ion charge control (ICC) defined target was set to 30,000, and the maximum accumulation time 200 ms. Collision induced dissociation (CID) MS/MS spectra were acquired for a *m/z* range between 50-2000, applying an isolation width of 2 Da and ICC target set to 40,000.

O*-*glycans were separated using the following gradient: 0-7 min, 1% B; 7-12 min, linear increase to 15% B; 12-70 min, linear increase to 40% B. Washing and re-equilibration steps were performed: 70-75 min, linear increase to 90% B; 75-80 min, held at 90% B; 80-82 min, return to 1% B; 82-90 min, re-equilibration at 1% B. The second valve switch was programmed for 83.5 min. For *O-*glycans analysis in IT-MS, MS spectra were obtained in negative ion mode between a mass range of *m/z* 450-1600, using UltraScan mode. The SPS defined target mass used was *m/z* 900, ICC defined target was 30,000, and the maximum accumulation time 250 ms. CID MS/MS spectra were acquired for a *m/z* range between 50-2000, applying an isolation width of 3 Da and ICC target set to 40,000.

Glycan structure determination and relative quantitation

Glycan composition was determined from the observed mass of each compound using GlycoMod(3), and MS/MS spectra were manually interpreted to elucidate glycan structure taking also the respective retention time of the compounds into account(2, 4-8). The software GlycoWorkBench (v2.1)(9) and UniCarb-DB (<https://unicarb-db.expasy.org/>)(10) were also used to aid in the glycan identification. The most relevant product ion signals present in the individual spectra used to assign a glycan structure are described in “Comment-Diagnostic ions (*m/z*)” (*Suppl Tables 4 and 5*).

Relative glycan quantitation was performed as described earlier using the area under the curve (AUC) from the respective extracted ion chromatograms (EICs) of the corresponding monoisotopic precursors(6, 7). Spectra visualisation and AUCs acquisition were performed manually using Compass Data Analysis v4.2 SR1 (Bruker, Bremen, Germany), with the integration limits being manually validated, after smoothing the chromatograms (Gauss algorithm, 1s, 1 cycle). Within the same sample, the AUC values of all the detected/quantified different glycans were summed (value corresponding to 100%) and the relative abundance determined for the corresponding AUC value.

Preview of the glycan data can be access on GlycoPOST(11) using the following credentials:

<URL:https://glycopost.glycosmos.org/preview/192552055960853e1fc17f0>

PIN CODE: 5775

#### High-pH fractionation of peptides

50 µg of protein (samples R7B1, R7B3, R7B6, BM0 and BM37) and 10 µg (samples R7B11 and BM41) were reduced, alkylated and precipitated with Chloroform-Methanol as described above. The protein pellet was air-dried for 10 minutes before 100 µL of 25 mM of ammonium bicarbonate (Sigma, Cat#09830) were added to the pellets. Trypsin was added at a ratio 1:25 (enzyme:protein ratio) and samples were incubated for 18 hours at 37°C. After digestion, trypsin was heat‑inactivated at 95°C for 10 minutes, and samples were dried under vacuum. 1000 U of PNGase F (2 µL) prepared in 50 µL of H_2_^18^O (Sigma, Cat#329878) were added, and samples were incubated at 37°Cfor 3 hours to deglycosylate N-glycopeptides before drying under vacuum.

Peptides were resuspended in 300 µL of 0.1% trifluoroacetic acid (TFA) and fractionated using a Pierce™ High pH Reversed-Phase Peptide Fractionation Kit (Sigma, Cat#84868) following the manufacturer’s instructions. Briefly, the resuspended peptides were loaded to the pre-conditioned supplied spin columns, and washed (3,000*xg*, 2 min) once using water. Increasing concentrations of acetonitrile (ACN) (5%, 7.5%, 10%, 12.5%, 15%, 17.5%, 20%, and 50%) in 0.1% triethylamine (TEA) buffer were used to elute (3,000*xg*, 2 min) the bound peptides into eight distinct fractions. All the resulting fractions were dried under vacuum and kept at -20°C until analysis. Samples were resuspended in 0.1% TFA and peptide amounts were quantified using a Thermo Scientific™ NanoDrop™ One/OneC Microvolume UV-Vis Spectrophotometer.

#### C18-LC-ESI-MS/MS analysis of peptides

Based on the NanoDrop quantitation, a volume corresponding to 600 ng of peptides were injected of each fraction. A MonoCap C18 Trap column (0.2 x 50 mm, 3 μm particle sizes, 100 Å pore sizes, GL Sciences, Cat#5020-10033) was used for pre-column enrichment and MonoCap C18 Nano Flow (0.1x150 mm, 3 μm particle sizes, 100 Å pore sizes, GL Sciences, Cat#5020-10151) was the separation column. A 120-minute LC method developing a gradient over 83.5 min was used for the separation of the peptides and subsequent column re‑equilibration. Solvent A consisted of 0.1% TFA, solvent B was 90%ACN/0.1% TFA. The flowrate was held at 0.5 µL/min on the separation column. Sample loading onto trap column was achieved within 6.5 min at 1% B and a flowrate of 6 µL/min. The valve switch bringing the precolumn in line with the separation column was initiated at 6.5 min and a separation gradient was started as follows: increase of solvent B from 1-10% over 2.5 min, followed by an increase from 10-30% over 51 min, then further 30-35% over 15 min, 35-45% over 5 min, 45-60% over 5 min and 60-90% over another 5 min. The separation column was then re-equilibrated using following gradient of solvent B: 90% over 4 min, 90-50% over 2 min, 50-40% over 2 min, 40% over 4 min, 40-1% over 3 min, 1% over 15 min.

The Orbitrap Fusion mass spectrometer was operated in positive polarity mode with the Ion Transfer Tube temperature maintained at 300°C The cycle time allowed for a full MS scan and subsequent MS/MS events was 3 sec. The full MS scan was performed at a 60K resolution in the Orbitrap with an automatic gain control (AGC) target value of 1x10⁶ (normalised AGC = 250%) for a scan range between *m/z* 600-2000. Charge states between 2-8 were allowed. Dynamic exclusion was enabled for 15 sec after 1 time, with +/-15 ppm. Precursors were prioritised based on their higher charge states as well as their higher *m/z* values. Before fragmentation, precursors were subjected to Quadrupole isolation using *m/z* 2 narrow isolation window, maximum injection time 54 ms. MS/MS scan first mass was defined as *m/z* 110. Higher-energy collisional dissociation (HCD) spectra were obtained in Profile mode at 30K resolution in the Orbitrap with an AGC target value of 5x10⁵ (normalised AGC=1000%), following stepped HCD (sHCD) fragmentation using 27, 30, 33% collision energies.

The mass spectrometry proteomics data can be accessed and reviewed on PRIDE using the following credentials

Username:

Password: FldtRJ8T

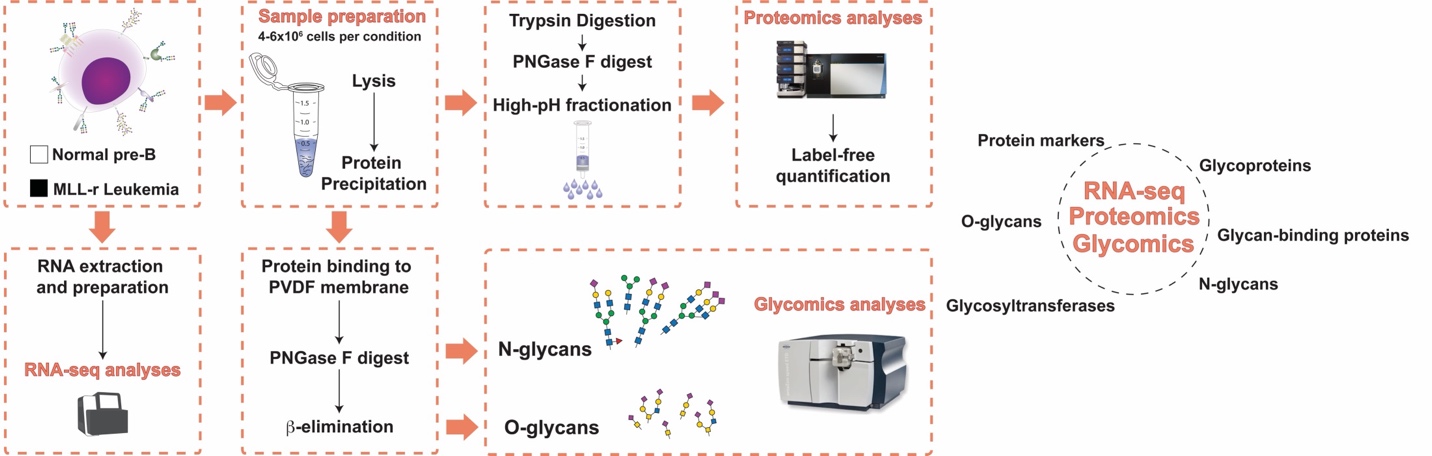

**Suppl. Fig. 1 Schematic of workflow of integrated glycomics analysis.**

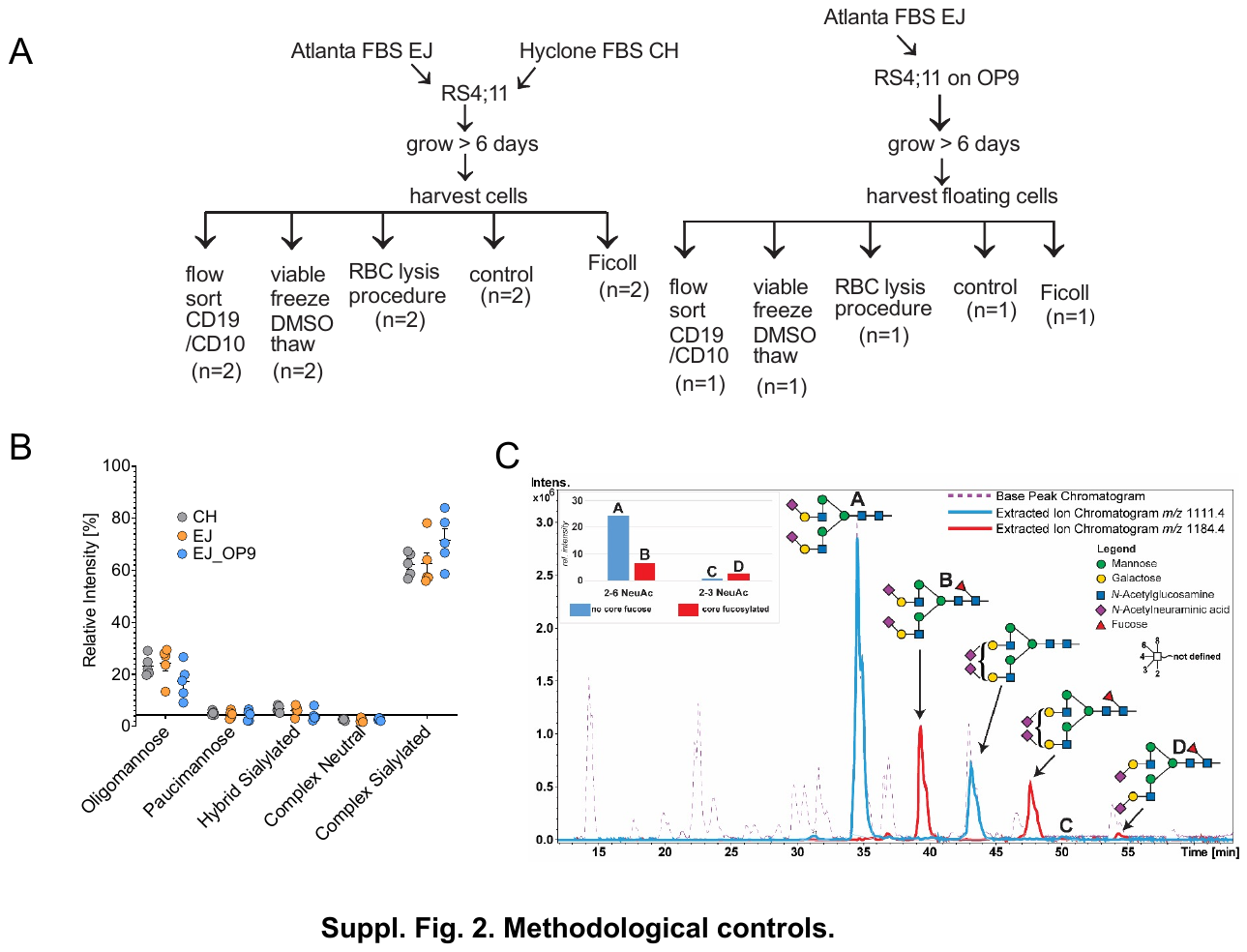

**Suppl. Fig. 2 Methodological controls.** (A) Schematic of RS4;11 cells subjected to different processing procedures including those used clinically on primary samples. (B) Relative intensities of glycans recovered in sample sets [EJ, CH] processed independently. OP9, samples co*-*cultured with OP9 stromal cells before glycan isolation (C) Analytical methods allow discrimination between closely related glycan structures.

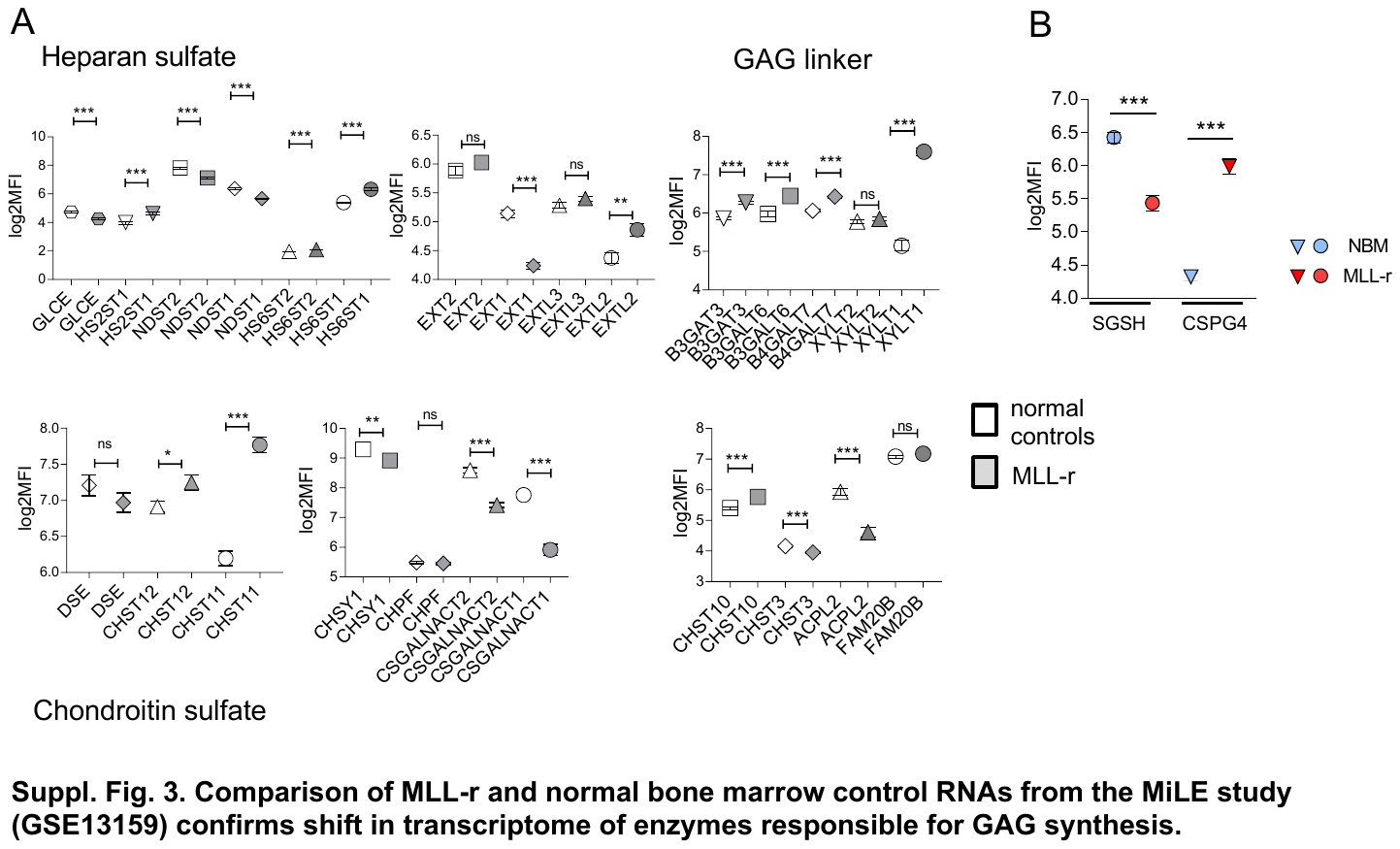

**Suppl. Fig. 3 Comparison of MLL-r and normal bone marrow control RNAs from the MiLE study (GSE13159) confirms shift in transcriptome of enzymes responsible for GAG synthesis.** (A) Transcriptome analysis using gene expression microarrays; n= 70 MLL-r and n =74 normal bone marrow control samples. All enzymes included in Fig. 5 are shown. (B) Expression of CSPG4 and SGSH. Values, mean ±SEM. ***p<0.05 as calculated by <http://servers.binf.ku.dk/bloodspot/>. The significance marks relies on t statistics for unequal sample sizes but assuming equal variance and the critical values are compared with a two*-*tailed probability. Graphs show the following probesets; XYLT1, 213725_x_at; XYLT2, 231550_at; B4GALT7, 53076_at; B3GALT6, 225733_at; B3GAT3 203452_at; FAM20B , 202916_s_at; ACPL2, 226925_at; CHST3, 208252_s_at; CHST10, 204065_at; EXTL2, 209537_at; EXTL3 209202_s_at; EXT1 201995_at; EXT2, 202012_s_at; HS6ST1 225263_at; HS6ST2, 1552767_a_at; NDST1 202607_at; NDST2, 203916_at; HS2ST1 203284_s_at; GLCE 213552_at; CSGALNACT1 219049_at; CSGALNACT2 239077_at; CHPF 202175_at; CHSY1, 203044_at; CHST11, 226372_at; CHST12, 222786_at; DSE 218854_at; SGSH, 35626_at; CSPG4 204736_s_at. Ns, not significant; *p<0.05; **p<0.01; ***p,0.001

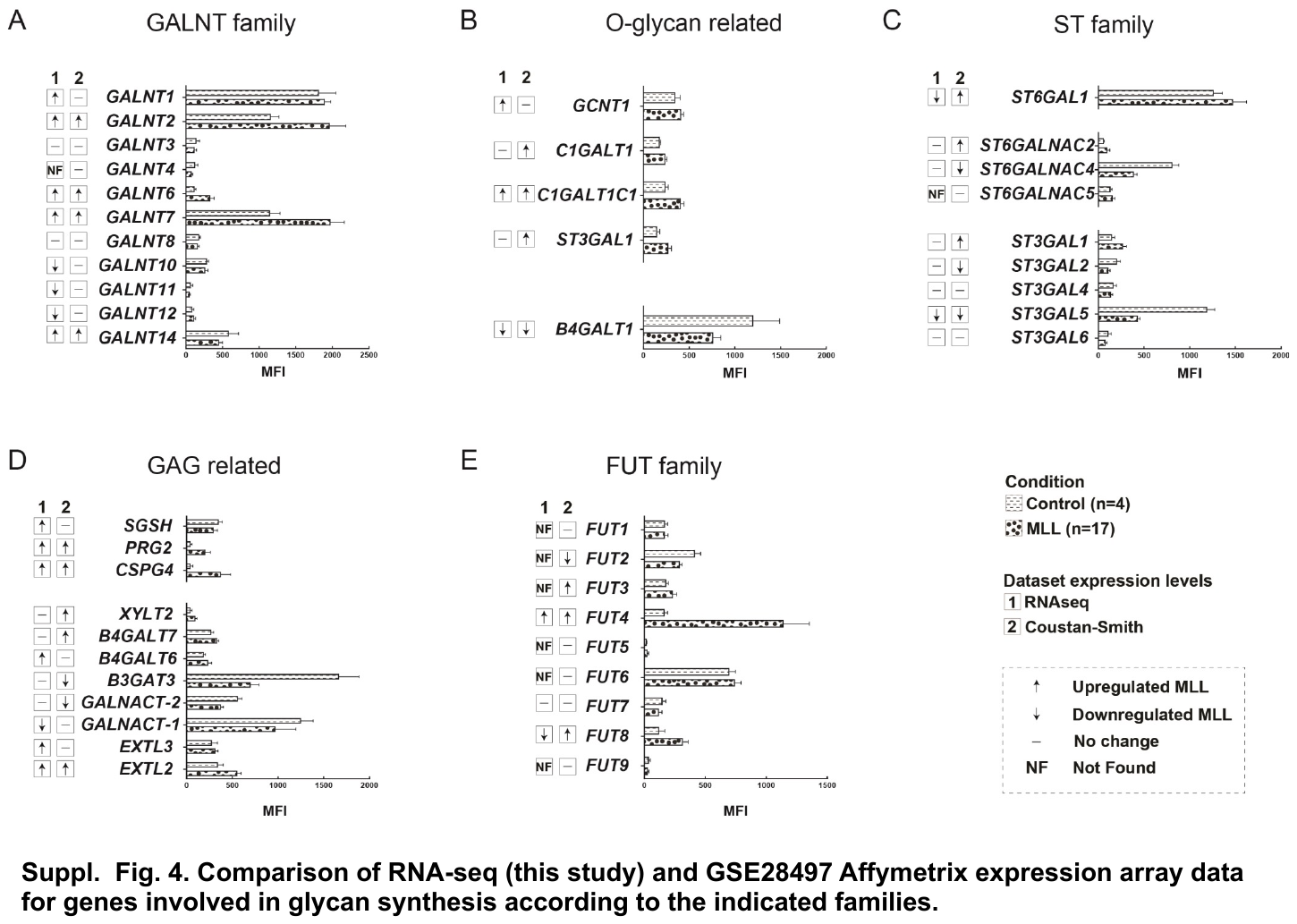

**Suppl. Fig. 4. Comparison of RNA-seq (this study) and GSE28497 Affymetrix expression array data for genes involved in glycan synthesis according to the indicated families.** Only data for the MLL-r and normal CD19+CD10+ bone marrow controls from Coustan-Smith *et al* (12) are included.

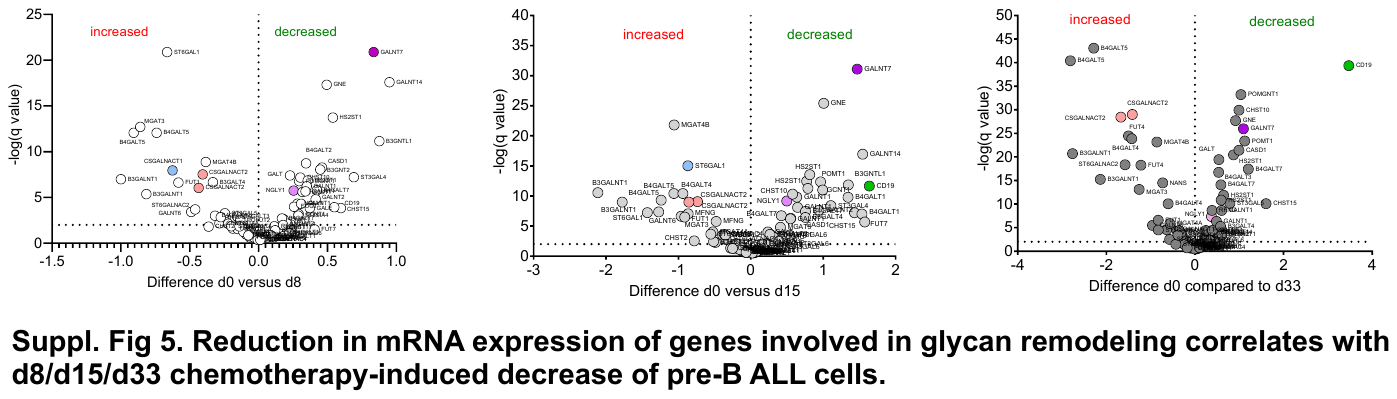

**Suppl. Fig. 5. Reduction in mRNA expression of genes involved in glycan remodelling correlates with d8/d15/d33 chemotherapy-induced decrease of pre-B ALL cells**. Meta analysis of RNA expression microarrays with 420 samples from 210 pediatric de novo ALLs at diagnosis, and d8, d15 and d30 of remission*-*induction therapy(13). Genes shown include those tested in the Cas9/CRISPR screen on 102 glycogenes; only genes of which there was significant differences in expression are shown. Not all genes were presented in this data set and some genes are represented by more than one probe set. *GALNT7*, purple circle; *CD19* [red circle] marks the demise of the leukemia cell population as a consequence of the chemotherapy.

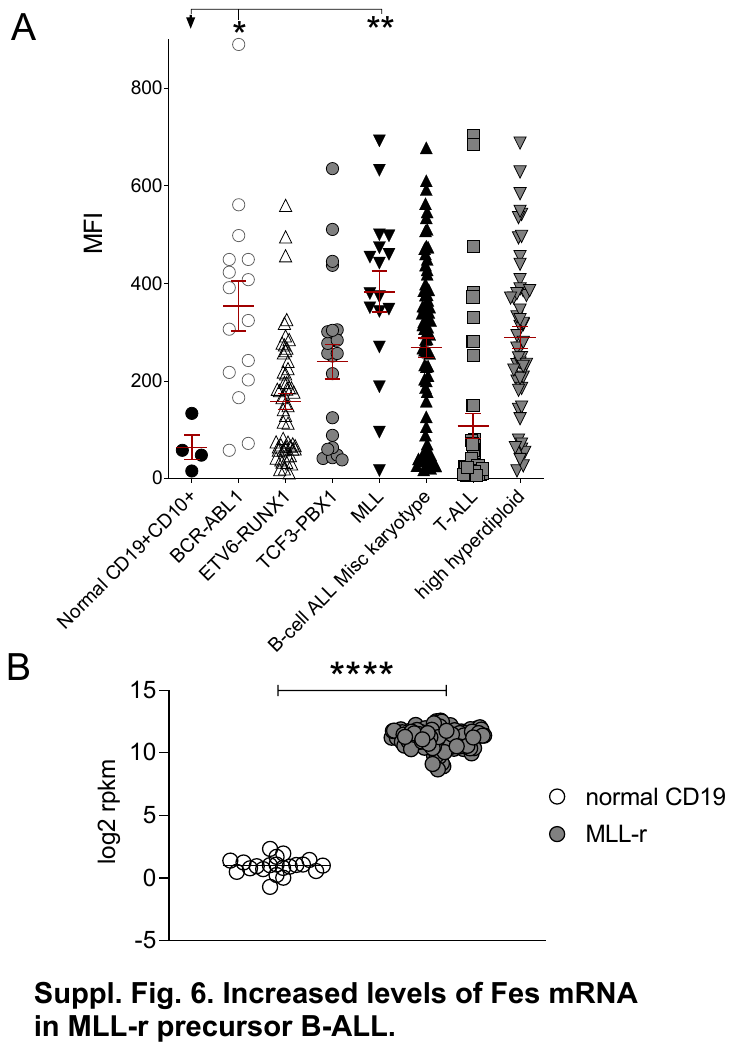

**Suppl. Fig. 6 Increased levels of Fes mRNA in MLL-r precursor B-ALL.** (A) GEO GSE28497 gene expression array data of 270 pre-B ALLs in different subcategories as indicated. MLL-r and BCR/ABL1: **p<0.01 and *p<0.05 respectively, compared to 4 normal bone marrow CD19+CD10+ precursor B-cell controls (B) RNA-seq [141 MLL-r and 21 normal CD19+ cord blood, peripheral blood and BM B-cells; Gu et al (14). ***p<0.01 ****p<0.001

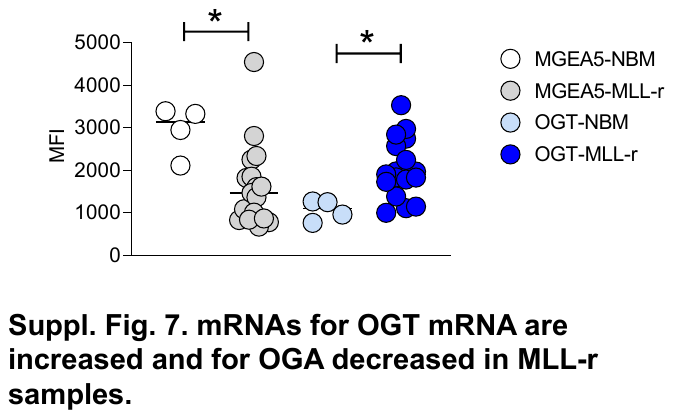

**Suppl. Fig. 7. mRNAs for OGT mRNA are increased and for OGA decreased in MLL-r samples.** GEO GSE28497 gene expression array data [17 MLL-r and 4 normal bone marrow (NBM) CD19+CD10+ precursor B-cell controls]. *p<0.05

**Suppl. Table 1 Summary of information of primary samples used for the integrated analyses**

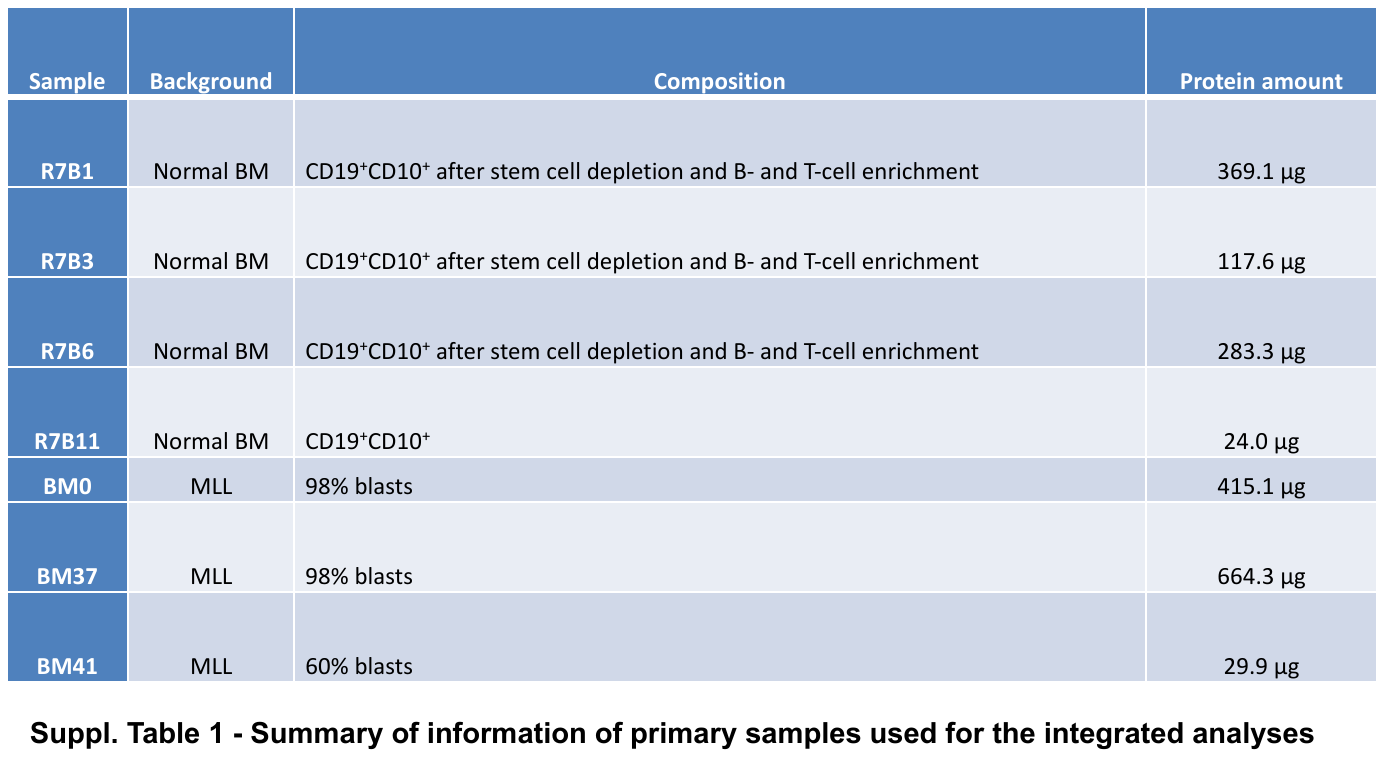

**Suppl. Table 2 Transcriptomics results**

**Suppl. Table 3 Proteomics results**

**Suppl. Table 4 O-glycomics results**

**Suppl. Table 5 N-glycomics results**

**Suppl. Table 6 Cas9/CRISPR screen: Impact of glycan*-*remodelling genes on KOPN8 cell survival**

**Suppl. Table 7 Transcript/proteomic integration**

### References

1.	D. Wessel, U. I. Flugge, A method for the quantitative recovery of protein in dilute solution in the presence of detergents and lipids. *Anal Biochem* **138**, 141-143 (1984).

2.	P. H. Jensen, N. G. Karlsson, D. Kolarich, N. H. Packer, Structural analysis of N- and O-glycans released from glycoproteins. *Nat Protoc* **7**, 1299-1310 (2012).

3.	C. A. Cooper, E. Gasteiger, N. H. Packer, GlycoMod--a software tool for determining glycosylation compositions from mass spectrometric data. *Proteomics* **1**, 340-349 (2001).

4.	A. V. Everest-Dass *et al.*, Comparative structural analysis of the glycosylation of salivary and buccal cell proteins: innate protection against infection by Candida albicans. *Glycobiology* **22**, 1465-1479 (2012).

5.	A. V. Everest-Dass, D. Kolarich, D. Pascovici, N. H. Packer, Blood group antigen expression is involved in C. albicans interaction with buccal epithelial cells. *Glycoconj J* **34**, 31-50 (2017).

6.	H. Hinneburg, F. Schirmeister, P. Korać, D. Kolarich, "N- and O-Glycomics from Minor Amounts of Formalin-Fixed, Paraffin-Embedded Tissue Samples" in High-Throughput Glycomics and Glycoproteomics: Methods and Protocols*,* G. Lauc, M. Wuhrer, Eds. (Springer New York, New York, NY, 2017), 10.1007/978-1-4939-6493-2_11, pp. 131-145.

7.	U. Moginger *et al.*, Alterations of the Human Skin N- and O-Glycome in Basal Cell Carcinoma and Squamous Cell Carcinoma. *Frontiers in oncology* **8**, 70 (2018).

8.	M. Pabst, J. S. Bondili, J. Stadlmann, L. Mach, F. Altmann, Mass + retention time = structure: a strategy for the analysis of N-glycans by carbon LC-ESI-MS and its application to fibrin N-glycans. *Analytical chemistry* **79**, 5051-5057 (2007).

9.	A. Ceroni *et al.*, GlycoWorkbench: a tool for the computer-assisted annotation of mass spectra of glycans. *Journal of proteome research* **7**, 1650-1659 (2008).

10.	M. P. Campbell *et al.*, Validation of the curation pipeline of UniCarb-DB: building a global glycan reference MS/MS repository. *Biochim Biophys Acta* **1844**, 108-116 (2014).

11.	Y. Watanabe, K. F. Aoki-Kinoshita, Y. Ishihama, S. Okuda, GlycoPOST realizes FAIR principles for glycomics mass spectrometry data. *Nucleic Acids Research* **49**, D1523-D1528 (2021).

12.	E. Coustan-Smith *et al.*, New markers for minimal residual disease detection in acute lymphoblastic leukemia. *Blood* **117**, 6267-6276 (2011).

13.	A. E. Yeoh *et al.*, Effective Response Metric: a novel tool to predict relapse in childhood acute lymphoblastic leukaemia using time-series gene expression profiling. *Br J Haematol* **181**, 653-663 (2018).

14.	Z. Gu *et al.*, PAX5-driven subtypes of B-progenitor acute lymphoblastic leukemia. *Nat Genet* **51**, 296-307 (2019).
